# Geospatial precision simulations of community confined human interactions during SARS-CoV-2 transmission reveals bimodal intervention outcomes

**DOI:** 10.1101/2020.05.03.20089235

**Authors:** B Goldenbogen, SO Adler, O Bodeit, JAH Wodke, A Korman, L Bonn, X Escalera-Fanjul, JEL Haffner, M Karnetzki, M Krantz, I Maintz, L Mallis, RU Moran Torres, H Prawitz, PS Segelitz, M Seeger, R Linding, E Klipp

## Abstract

Infectious disease outbreaks challenge societies by creating dynamic stochastic infection networks between human individuals in geospatial and demographical contexts. Minimizing human and socioeconomic costs of SARS-CoV-2 and future global pandemics requires data-driven and context-specific integrative modeling of detection-tracing, healthcare, and non-pharmaceutical interventions for decision-processes and reopening strategies. Traditional population-based epidemiological models cannot simulate temporal infection dynamics for individual human behavior in specific geolocations. We present an integrated geolocalized and demographically referenced spatio-temporal stochastic network- and agent-based model of COVID-19 dynamics for human encounters in real-world communities. Simulating intervention scenarios, we quantify effects of protection and identify the importance of early introduction of test-trace measures. Critically, we observe bimodality in SARS-CoV-2 infection dynamics so that the outcome of reopening can flip between good and poor outcomes stochastically. Furthermore, intervention effectiveness depends on strict execution and temporal control i.e. leaks can prevent successful outcomes. Schools are in many scenarios hubs for transmission, reopening scenarios are impacted by infection chain stochasticity and subsequent outbreaks do not always occur. This generalizable geospatial and individualized methodology is unique in precision and specificity compared to prior COVID-19 models [6, 16, 17, 19] and is applicable to scientifically guided decision processes for communities worldwide.

## Main

As the SARS-CoV-2 pandemic is spreading around the world it is inflicting multi-dimensional damage to humanity: millions of COVID-19 cases are bringing healthcare systems close to collapse, halting or suppressing global and local economies, and normal human life. In response, countries and communities are scrambling to fight the virus with a series of diffrent interventions (mitigation measures) and strategies aimed at preventing new infections whilst providing optimal treatment of patients and aiming to reopen societies and economies as quickly as possible. To assist governments in choosing between different intervention and reopening strategies, data-driven mathematical and computational modeling can predict the trajectory and severity of infection outbreaks, the expected number of fatalities and the effect of different interventions.

Considering the lack of information about the disease (COVID-19) but also its near- and long-term impact on healthcare systems and societies, precise and context specific models are required for deciding optimal strategies for reopening and allowing populations to return to work and social life. To understand transmission of SARS-CoV-2/COVID-19 and to guide such decision processes in a geolocation-specific manner, detailed simulation of prolonged or intermittent patterns of social/physical distancing is required in order to prevent healthcare systems and communities from collapsing. Therefore, it’s essential to capture the stochastic nature of individual transmission events. Traditional epidemiological/statistical models cannot make predictions in a geospatial temporal manner based on human individuals in a community and cannot for example capture local particularities. Thus, the challenge is to conduct spatio-temporal simulations of transmission networks with real-world geospatial and georeferenced information of the dynamics of the transmission and disease progression and the effect of different intervention strategies such as isolation of infected individuals or location closures.

Here, we present a comprehensive agent-based model that incorporates the clinically described stages of SARS-CoV-2 infection, COVID-19 disease and recovery. It incorporates demographic data, realistic daily schedules and, importantly, the *physical location* of individuals. We comparatively simulate and analyze different tailored scenarios and non-pharmaceutical interventions, depending on the location (e.g., workplace, school, public places such as shopping malls, etc.) but also on the actual time of day. To this end, we have integrated a large amount of publicly available data, e.g., on locations, age distribution, household composition, daily occupation and schedule, geographical information, and sociological data for typical numbers and types of social contacts in the population.

By means of spatial, georeferenced and demographic stochastic modelling of COVID-19 infection networks, we present new insights into the pandemic and decision support for (non-pharmaceutical) intervention and exit strategies.

## Results

### An individualized GEoReferenced Demographic Agent-based model (GERDA)

We model the virus propagation and effects of non-pharmaceutical interventions within a concrete population (**Fig. 1**). The model tracks a number of individuals with specific infection states at specific physical locations over time. Due to its geospatial design, GERDA makes use of the actual number of residential buildings, workplaces, schools or public places in a given community as the space for potential infection chain initiation by human contact. As an example, we used data from the municipalities of Gangelt and Heinsberg (**Supplementary Fig. 2**), which experienced significant and early COVID-19 outbreaks. Gangelt had more than 300 confirmed cases among 12,000 inhabitants with the outbreak after a carnival event in March 2020 followed by widespread transmission within the community. This outbreak is one of the best-characterized worldwide with comprehensive epidemiological monitoring and sampling, which renders this municipality a solid starting point for developing GERDA.

**Fig. 1:**
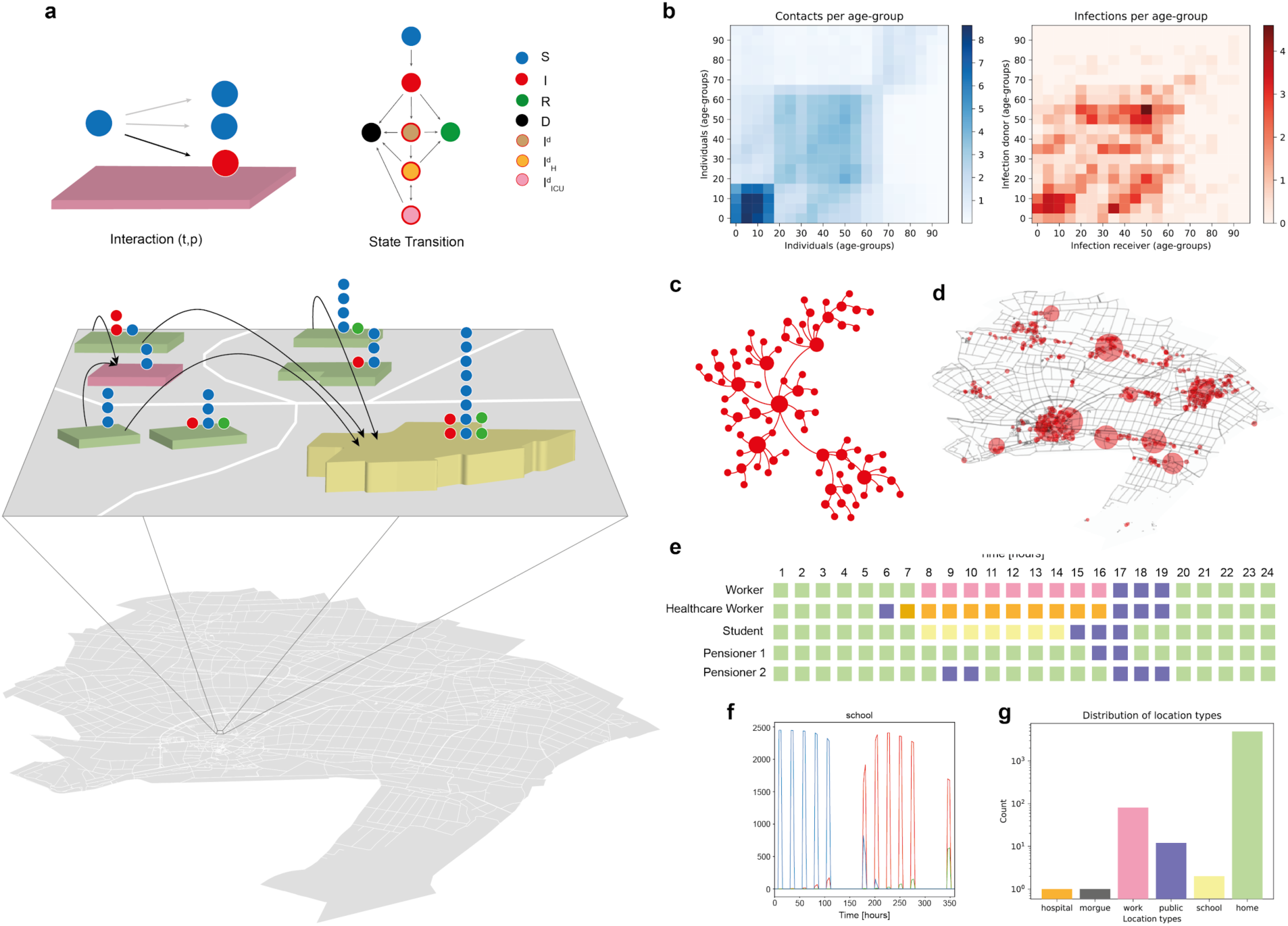
Overview of the geospatial, individualized demographic and georeferenced model (GERDA) **a** The model encompasses a real-world community (bottom), exemplified by a small-town municipality (Gangelt, Germany). The world is created from coordinates and building type information. The human individuals (represented by computational agents) each have individual properties such as age, infection state, and occupation drawn from census-based demography. Each individual inhabits a location at any point in time, travels between locations based on specific weekly schedules and can encounter other individuals (**e**), thus creating an interaction network through which the infection can spread. The individual’s health status changes via location-dependent contacts with infected individuals and data-based transition probabilities, allowing state transitions between susceptible (S), infected (I), recovered (R), deceased (D) and infection sub-states diagnosed (I^d^), hospitalized (I^d^_H_), in intensive care (I^d^_ICU_). **b**, Average number of daily unique contacts (left) and average number of infections per day (right), between members of different age groups in the baseline scenario over a period of 5 working days. **c**, Representative infection chain network, originating from one infectee. **d**, Snapshot (from Movie 1) of infection dynamics at a specific time point, showing the number of infected individuals (circle size) at different locations in the community. **f**, Hourly occupation of the school, based on the schedules, over the course of two weeks, lambed by the specific statii (S: blue, I: red, R:blue). Note the change in status on the second Monday (168h) indicating infection in school. **g**, Number of locations of different types in the modeled town.

In the model a human being is described as an autonomous agent who is always present at one specific physical location at any given time point. The location can be one of the following types: residential buildings (home), work places, schools, hospitals, and public places; deceased individuals are assigned to a morgue location. These *locations* are initialized automatically from openly available data for a given municipality using geospatial data from OpenStreetMap (Methods). The population is initialized with demographic data specific to the geolocation (municipality) from the relevant census resulting in representative age distributions and household compositions. Every individual has three features: an age (in years), a weekly schedule resolved by the hour, and a health status. The weekly schedule determines an individual’s presence in each of the possible location types at each hour. Schedules can change with health state and interventions. The health status for individuals are: susceptible (S), infected (I), recovered (R) or dead (D). Infected individuals (I) can obtain sub-states specifying their condition as diagnosed (I^d^), hospitalized (I^d^_H_), or being in an ICU (I^d^_ICU_).

The model is initiated with a basic initialization of households, workplaces, public places, schools, a hospital, and a morgue as well as a total number of infected individuals. The latter originates from assigning a household (randomly drawn from the demographic distribution) to each residential building.

During the simulation, agents visit locations specified by their respective schedules and the infection spreads across the spatio-temporal network of interacting agents, resulting in a contact pattern (**Fig. 1b** and **Supplementary Fig. 11**) resembling the data by Zhang et al [17]. This spatio-temporal network, defined by periodically recurring movement patterns, constitutes the environment in which the agents interact and the infection spreads. Agents operate in defined sub-networks, specified by regularly visited locations.

### Absence of interventions leads to an uncontrolled outbreak trajectory

The baseline scenario simulates the trajectory of infection progression in absence of any interventions and provides a benchmark (spread of both diagnosed and undiagnosed infection) for evaluating subsequently introduced interventions (**Fig. 2**). The time courses for states (S), (I), (R) and (D) as well as substates (I^d^), (I^d^_H_), and (I^d^_ICU_) are simulated with 100 repetitions (**Fig. 2a,b**); age-dependent trajectories are also available (**Supplementary Fig. 1**). Reflecting the progression of the outbreak, the regular visits to different locations by individuals within the community (e.g. public places predominantly during weekends and workplaces/schools during weekdays) changes towards diagnosed citizens staying at home or in the hospital (**Fig. 2c,d**). An increasing number of deceased individuals is assigned to the morgue.

**Fig. 2:**
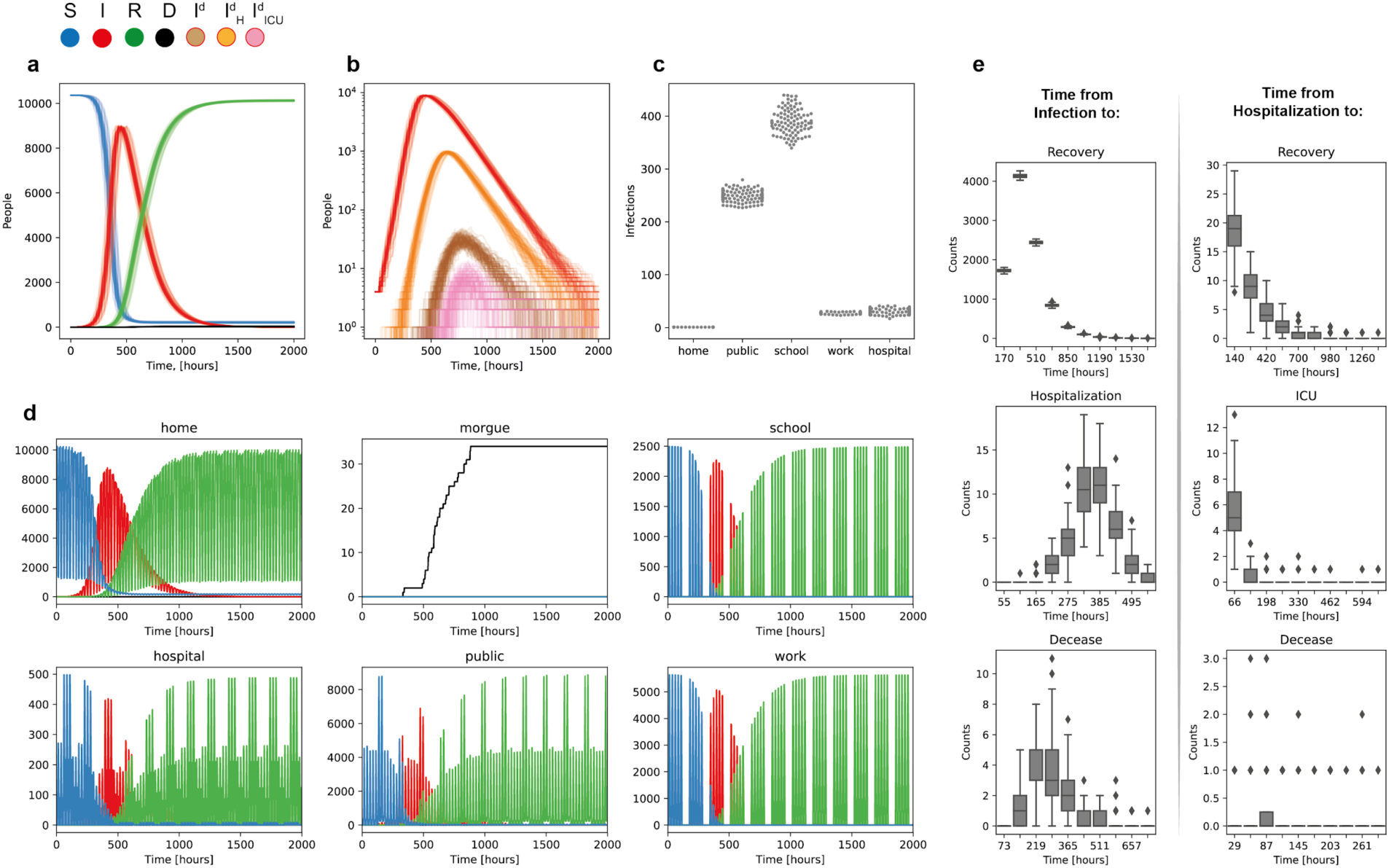
The baseline scenario. **a** Trajectories of different status groups (S), (I), (R), and (D) resulting from 100 simulations. **b**, Trajectories of the corresponding infection sub-states (I^d^), (I^d^_H_) or (I^d^_ICU_). **c**, Total infection events per instance for different location types over the full simulation period of 2000h (approximately 83 days). **c**, Hourly resolved occupation of location types by different status groups. Weekday/weekend and night-/day shift specific rhythms are apparent from e.g. the hospital panel. **e**, Distribution of status dwelling times before transiting to a specific (sub-) status for 100 simulations. Parameter value: Infectivity *k_inf_ = 0.3*.

The model parameters are based on published data for transition periods between infection and diagnosis, diagnosis and hospitalization, frequencies of need for ICU as well as death rates (**Methods**). In order to verify that the model simulations reflect these underlying data in a satisfactory manner, we determined distributions of transition frequencies per unit of time for the different state transitions considered in our model (**Fig. 2e**). These frequencies agree well with the frequencies reported in literature, although not all necessary values for all age groups have been published yet.

Deploying GERDA we simulated the infection spreading in two German communities (Gangelt and Heinsberg), a British (Epping) and a Swedish community (Vaxholm near Stockholm). Simulations show that in all four communities, major infection transmission hubs are locations at which people from different ages and subgroups meet (schools in Gangelt, public places in Heinsberg and Vaxholm; **Fig. 2** and **Supplementary Fig. 2**), rendering closure of these a likely effective way to interrupt infection chains. One possible explanation is the size difference between the two, i.e., e.g. having more schools decreases the impact of each of them, rendering public spaces that allow interactions between diverse otherwise completely isolated subgroups more influential and important for infection transmission. Even though this hypothesis warrants further studies, the fact that closure of one location type in one municipality might sufficiently interrupt infection chains while in another it might not prevent the infection wave showcases the criticality of real-world data for informed decision making.

The temporal and spatial dynamics of infection spreading resulting from the simulation of the baseline scenario is visualized in Movie 1 [https://www.tbp-klipp.science/GERDA/Movie-1.mp4] with a still image in **Fig. 1**. It is important to note that the daily rhythm shown in the movie results from individuals moving between their respective homes and workplaces. The infection starts with two infected individuals in one household at time 0 h. At times 100 h and 200 h more and more infected agents are observed, especially at the geospatial hubs (e.g., center of the town).

### Effectiveness of non-pharmaceutical interventions depends on duration and stringency

In order to analyze the effect of interventions such as lockdowns or contact prohibition, we tested a series of scenarios (**Fig. 3**): (**i**) Scaling of the infectivity as proxy for wearing face masks or keeping physical distance, (**ii**) Selective closure of public spaces such as schools, general public spaces, workplaces or a combination of these, and (**iii**) Effect of influx of infected people.

**Fig. 3:**
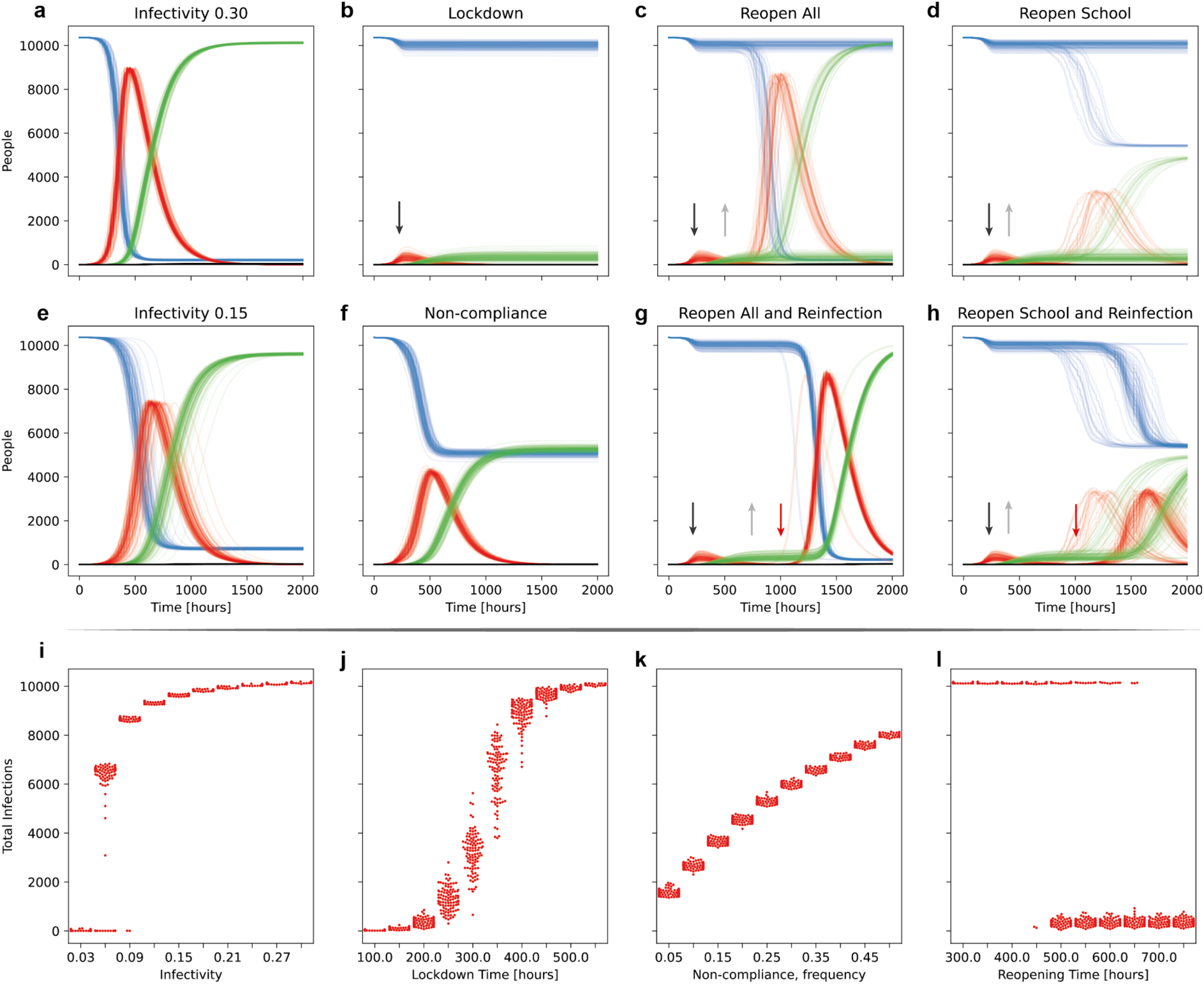
Effect of non-pharmaceutical intervention scenarios, and individual behavior on the time course of infection. **a**, Baseline scenario with infectivity *k_inf_ = 0.3*. **b**, Scenario with full lockdown at time 200 h (about 16 days, indicated by downward arrow). Work, school and public are closed and individuals have to stay at home instead. **c**, Reopening of all locations at time 500h (about 20 days, indicated by grey upward arrow) after lockdown as in **b. d**, Selective reopening of only schools at 450h after lockdown as in **b. e**, Reduction of infectivity mimicking wearing face masks or keeping physical distance. **f**, Lockdown as in **b**, but 25% of individuals can or will not comply. **g**, Lockdown and reopening all at 750h, with entry from outside of the community of 5 new infected individuals at time 1000h (about 40 days, red arrow). **h**, Lockdown and reopening of schools as in d with 5 newly entered infected individuals at 1000h. **i-l**, Distribution of the total number of infections in 100 simulations for different parameter values of the model: **i**, Variation of infectivity, **j**, Variation of the lockdown time as in **b, k**, Variation of the non-compliance frequency as in **f, I**, Variation of the reopening time of all locations as in **c**.

Here, we have compared the baseline scenario with infectivity values of *k_inf_=*0.30 (indicating typical behavior) and values lower than 0.3 corresponding to different levels of social distancing (**Supplementary Fig. 3**). As example, *k_inf_*=0.15 means the probability to get infected when meeting an infected individual is decreased by 50% (**Fig. 3e**). Reduced infectivity leads to a slower infection wave with a lower and later peak of (I). Ultimately, fewer people are infected and move to the recovered state because the overshoot in infected individuals is reduced [5]. Variation of infectivity between 0.03 and 0.3 leads to a pronounced jump in the total number of infections at a value of about 0.06 (**Fig. 3i**).

More restrictive interventions to influence the course of the epidemic analyzed with GERDA include closure of all locations except of homes and hospitals (i.e., workplaces, schools and public spaces) as well as reopening of all or selected locations or closure of selected locations (**Fig. 3**, 100 simulations each). Closure of all locations after 200 h (ca. 8 days) leads to significant reduction of the infection peak and final number of recovered individuals. However, the impact of measures strongly depends on early closure (**Fig. 3j** and **Supplementary Fig. 4** and **15**). Closing just a few days too late significantly increases the mortality, as has been described for example in the United Kingdom. Subsequent reopening of either schools, public or workplaces at about 500 h (ca. 20 days) results in a unique phenomenon, namely a bimodal behavior of the system. In this case, the infection ceases in some simulations, while generating a strong second peak in others (**Fig. 3l** and **Supplementary Fig. 5** and **6** for variation of reopening time points).

**Fig. 4:**
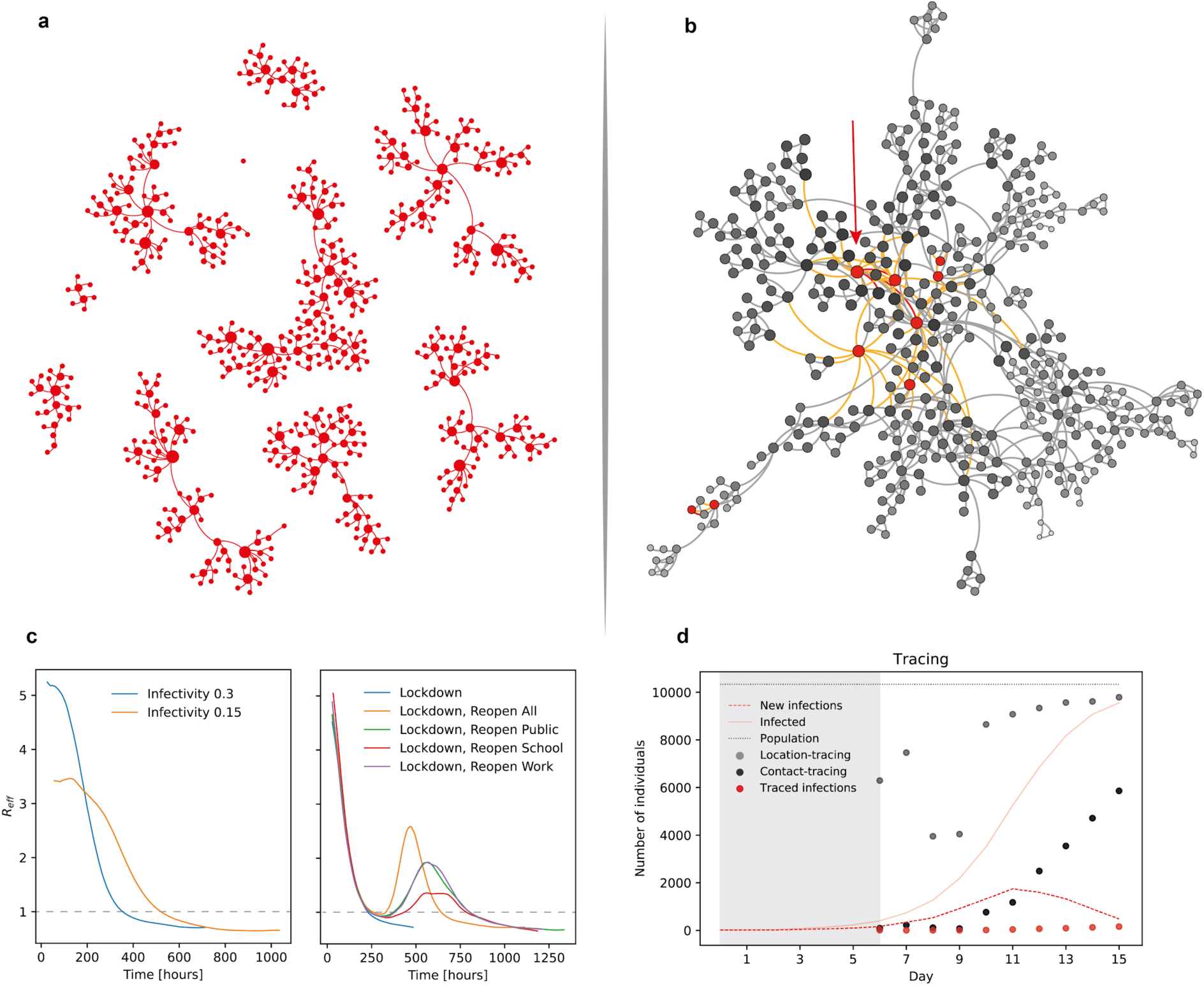
Infection network based tracing. **a**, Network of infections starting from 10 infected individuals leading to an unconnected network (baseline scenario; snapshot after 200h to retain visibility). **b**, Temporal network of possible infection transmissions starting from one individual (source, red arrow) unfolding within 15h after the source creates the first subsequent infection. Red edges: interactions leading to new infections, orange edges: interactions with non-realized potential for infection, grey edges: all other interactions without potential for infection. Red dots: infected individuals (from this or other sources), grey dots: non-infected individuals, grey-scale indicates distance to infection source (number of contacts in-between). **c**, R_eff_ values for the baseline scenarios and different scenarios of non-pharmaceutical interventions. **d**, Evaluation of the efficacy of location- and contact based infection tracing. The first diagnosed individual appears at day 6 and the number of individuals, identified by back-tracing over the previous 10 days, are indicated for contact-based (black dots) and for location-based (grey dots) tracing. Red dots indicate the number of infections by the diagnosed individual, during the last ten days, which would be diagnosed when tested. An indication of the total number of (cumulative) infections is given by the solid red line and the total infections per day are represented by the dashed red line. For orientation, the population size is indicated by the dotted grey line. Note that in real world tracing-based testing would alter the infection, diagnosis and ultimately subsequent tracing dynamics. The simulation only represents the case, when tracing based testing would not have been utilized before and, thus, represents the information at the onset of tracing-based testing.

In contrast to the lockdown time, the total number of infections does not depend gradually but in bimodal fashion on the time of reopening, i.e. is either high or low with increasing probability of low infections for later reopening times (**Fig. 3l** and **Supplementary Fig. 12**). This is due to the stochastic nature of the process: the precise behavior of individuals is unpredictable, despite being regulated. The ratio of cases of declining and recurring infections varies between the scenarios, as well as the height of the second infection peak with the strongest peak when all public activities are resumed in the scenario “reopen all”. However, already selective opening of only schools bears a strong risk of a second infection outbreak. We also investigated what happens if a number of individuals will not follow the order for closure of all locations. This reflects either non-compliance (e.g. civil disobedience) or the fact that some people in systemically relevant jobs have to go to work or need to send their children to school (or daycare) to be able to go to work themselves. We see that already small levels of non-compliance have severe effects, i.e. 10% or 25% non-compliance would lead to a severe rise of infected and later recovered individuals (**Fig. 3f,k** and **Supplementary Fig. 7** and **13**).

Simulating the emergence of newly infected individuals from outside, e.g. due to travel or visits, a second infection peak is very likely as long as there is still a pool of susceptible individuals (**Fig. 3g,h** and **Supplementary Fig. 8**).

In summary, the only perfect effective scenario to prevent a second outbreak is sufficiently long extension of full lockdown until no infectious individuals remain. These results have serious consequences for considering the reopening of societies and communities: In our simulations, early reopening of schools or of all locations can lead to a strong second peak of infection, while reopening after sufficiently long closure is safe as long as no new infected individuals enter the community.

### Non-pharmaceutical interventions such as location closure reduce the effective reproduction number R_eff_

The GERDA model can be used to monitor the effective reproduction number R_eff_ in a time-dependent fashion, as an emergent property of the modelled system (**Fig. 4**). In the baseline scenario with an infection rate *k_inf_ = 0.3* the value ranges between about 5.2 at the beginning of the infection wave and 0.71 at the end of the simulation period. When we reduce the infection rate to *k_inf_ = 0.15* mimicking medium social distancing, the values range between 3.5 and 0.66. Lockdown of schools, public and work leads to faster decline in R_eff_. The reopening of all or selected locations induces a second increase in R_eff_, the fastest for reopening of all locations. All lockdown and reopen scenarios ultimately end with R_eff_ values of about 0.7.

### Network based benchmarking of tracing strategies

Relationships between individual agents in GERDA constitute a network of hierarchical edge-types, which can be reconstructed from the simulation (Fig. 4). The basal and most generic network is represented by the spatio-temporal overlap of agents visiting the same location at the same time (co-location network). The network of interactions between agents constitutes the second layer and is a sub-network of the co-location network. It comprises three different types of interactions, namely those that: (**i**) cannot lead to infection (none of the interaction partners is infectious), (**ii**) can potentially result in transmission, but do not (interaction between (S) and (I) without successful transmission), and (**iii**) result in transmission of infection from (I) to (S) (**Fig. 4a, b**).

Inspired by recent efforts to develop tracing apps and in order to further elucidate the potential of this approach, we tested the efficacy of co-location and contact-based tracing methods, in particular at the onset of their application. Co-location tracing is realized by identifying individuals’ spatio-temporal overlap, contact-tracing by identifying the actual interactions (potentially facilitating infection transmission). Upon diagnosis of an infected individual, the (potential) transmission targets for the preceding 10 days are identified. The number of identified individuals differs vastly between the two methods, while the infection-transmitting contacts are a subset of both of them. Thus, the expected proportion of positive results, when testing all traced contacts, is 20 times higher for the contact-based, than for the location-based tracing (with on average 50 vs 1000 required tests per identified case).

The high dark figure of non-identified infectees leads to an average fraction of 15-20% of infections identifiable by tracing. Consequently, this identification rate will increase when incorporating recursive tracing into the model. Thus, otherwise hidden infectees are identified and subsequently isolated, resulting in early interruption of infection chains.

In the baseline scenario, the number of traced individuals rapidly approaches a significant proportion of the population (**Fig. 4d**), a process that is slowed if the infection rate is lowered by interventions. A timely adoption of contact tracing and collection of the required tracking data, prior to the first diagnosed case; are key for full exploitation of the short time frame with advantage over undifferentiated mass testing. Though the power of this approach can be further explored, our results suggest that cell phone-based contact tracing might also serve for pre-emptive sentinel monitoring during lockdowns or an effective control measure accompanying reopening (**Supplementary Fig. 9**).

## Discussion

By creating a real-world data-derived environment, GERDA simulates the SARS-CoV-2 infection spread and the corresponding COVID-19 societal burden by explicitly integrate interactions between human individuals, individual infection events, and human demographic characteristics (household composition, occupation, daily movement patterns, age and disease state) and physical proximity networks. This approach renders it possible to create a model, based on realistic assumptions and microscopic insights into the mechanics of infection spread. It enables GERDA to model realistic chains of infection and targets for intervention measures in georeferenced environments.

Classical SIR-models [e.g.,1,2] are based on population-wide assertion, thereby omitting the variability of the infection transmission between individuals, neglecting individual behavior and geospatial data and being not suitable for real-time simulation. Traditional agent-based models [e.g., 3] capture stochasticity in agent behavior, but are highly limited in size by computational complexity nor cover specific geolocations. Others have attempted to overcome these limitations: Already back in 2005, Ferguson et al. [18] published an agent-based model considering statistical distributions of transmission events in households, building types and alike. More recently, Gomez et al. [20] simulated infection transmission throughout Bogotá representing the city’s population by 1000 agents while Lai et al. [19] created a simple model to analyze the effect of traveling between cities in China, but without realistic individual agents. Still, they all attempt to describe the infection process on population level, thus relying on a coarse-grained representation by a limited number of agents. Alternatively, Kissler et al. [6] developed an ODE model that describes increased social distancing by decreasing the *R* value. Ferretti et al. [16] modeled pre-symptomatic and non-diagnosed transmissions whereas Zhang et al. [17] provided valuable age specific data on contact patterns in the Chinese population. Karin et al. [21] assessed the effect of alternative working schedules. Certainly, all of those models can provide useful information for decision processes during a pandemic, however, they all focus on population dynamics and can neither consider geospatial referenced motion of agents in their communities into account, nor individually varying behavior, nor analyze infection spreading and disease progression on an individual scale.

In contrast, our approach aims for precision: it explicitly considers individual characteristics, the physical structure of a society described by geolocations and inhabitant patterns in the created environment. Simulations create a dynamic interaction network that due to complete time course tracking allows to compare contact patterns for the baseline and intervention scenarios. This “microscopic” approach is computationally more expensive but has several advantages compared to other epidemiological models: (**i**) it is able to perform comparative analysis for different intervention scenarios based on their relative effectiveness. (**ii**) it can deploy interaction networks between human individuals to evaluate and benchmark different testing and tracing strategies. (**iii**) the effective reproduction number *R* is an emergent property of our model, not an *a priori* fixed parameter, thus providing a direct measure for the efficacy of intervention strategies. (**iv**) it can accurately model localized and smaller outbreaks as a basis for analysis of the dynamics and optimal strategies not accessible by statistical models.

We demonstrated these capabilities by deploying GERDA on two small-towns (Gangelt and Epping) and larger-town (Heinsberg), neighboring communities, as well as a Swedish community (Vaxholm near Stockholm), simulating selected intervention and reopening scenarios and comparing to a (worst case) baseline scenario of no interventions. For example, simulations for Gangelt with a lockdown 8 days after the suspected initial transmission event and with 10 % non-compliance match the levels of infections and deaths reported for Gangelt with a high degree of accuracy (**Supplementary Fig. 10**).

Importantly, the predicted outcome for reopening scenarios reflects the stochasticity of infection events i.e. in the majority but not in all cases a subsequent outbreak takes place (**Fig. 3**). This bimodality demonstrates that effective interventions require strict execution (stringency) and careful temporal control (timing). This is also supported by the observation that closing just a few locations while confronted with a high proportion of non-compliant individuals, in many scenarios does not qualitatively change the simulation output compared to the baseline scenario. The bimodality, however, also implies that early exit from lockdown does not necessarily lead to a second outbreak, which we observe across different simulated German, British, and Swedish communities (**Supplementary Fig. 12**).

A general limiting factor in disease modelling, which also holds true for GERDA, is that the transition probabilities for the propagation of e.g. COVID-19 are intrinsically incomplete and evolving (**Supplementary Table 1**, [4]). Another caveat in our study is the setting of four relatively smaller european communities with a limited number of schools, work- and public places. Currently, GERDA does not include potential COVID-19 “hot spots” such as kindergartens, public transport networks, universities or nursery/senior homes, the latter of which can contribute up to 50% of mortalities. They will be incorporated in future versions. Given different dynamics might be observed when simulating densely populated or connected areas of megacities, metropoles and suburbs, future studies of larger and more “complex” communities with more inhabitants, location types, and travel activities are warranted. Therefore, the long-term goal is to extend GERDA to cover diverse and metropolitan areas, encompassing entire countries, regions and the globe in a community effort.

Important for public health recommendations based on cell-phone tracking is that, whilst the network created by GERDA can elucidate the effect of contact tracing and testing strategies, its predictive capacity could be improved by including feedback mechanisms from tracing of diagnosed individuals to the modeled diagnosis process.

Remarkably, simulating these relatively small communities GERDA is the first model to offer insight into the bimodal behavior of SARS-CoV-2 infection dynamics and to facilitate comparative analysis of intervention strategies in a specific community. With unprecedented level of detail, GERDA provides novel insight into the different interaction networks facilitating the underlying infection propagation in a human population. Furthermore, this makes it possible to analyze infection chains as precisely as in reality possible only with virus genome sequencing in addition to testing the entire population, but also to benchmark the effectiveness of tracing and testing strategies.

The COVID-19 pandemic is hurting the wellbeing of children across many communities and as a consequence there are ongoing debates on reopening of schools in many countries. Unfortunately, our results show that this in many (though not all scenarios) can trigger a major subsequent infection wave. We propose that a direct evaluation of these consequences must be conducted, given children must be protected independent of the impact on adults.

Despite our work being motivated by the 2020 corona disaster, the concepts and planned extensions will be valuable when responding to e.g. the common flu or other pathogens. Furthermore, in the future it could be groundbreaking to extend the model to global communities and include modeling of therapeutic and vaccine interventions in order to direct public health policies and better safeguard the public preferably in real-time.

## Methods

The ABM simulation framework was designed in an object-oriented manner, using the programming language **Python** version 3 [7] and the packages **NumPy** [8], **pandas** [9], **GeoPandas** [10], **OSMnx** [11]. The packages **Matplotlib** [12], **seaborn** [13] and the software **gephi** [14] were used for visualization. Despite being streamlined for the parallel execution of numerous replicates; the developed tool was designed for usage on customary computers.

### Design of the GERDA Model

The model is agent-based and georeferenced, thus it comprises a world and agents. Each agent represents an individual, occupying a specific location at each time-step.

#### Characterization of the world

The modelled world represents the chosen area, based on the available geo-data. It is defined by the coordinates and types of various locations or buildings, where the distances between locations affect the agents’ movement-patterns. The current version distinguishes between the location-types residential/home, work, school, hospital and public/leisure.

#### Characterization of the agents

Agents have *three main features:* age, (infection) status, and schedule. Age and status can be permuted without limits, however the choice of schedules is restricted by age and status.

The *age* of an agent lies between 0 years and 99 years. It is randomly drawn from a distribution based on household compositions, resembling, e.g., German demography, according to census (**Supplementary Fig. 14**). Agents are characterized according to their health *status* as susceptible (S), infected (I), recovered (R) or deceased (D). Agents among the group of infected individuals (I) obtain sub-states specifying their condition as (only) infected (I), diagnosed (I^d^), hospitalized (I^d^_H_), or being in an ICU (I^d^_ICU_). Each agent has a *schedule* containing the locations visited by the agent with resolution of an hour, including discrimination of weekdays and weekends, while every day can be different. Distinct types of schedules pertain to the groups of children/students, adults/employees, adults/pensioners. Schedules change depending on (i) the status of the agent or (ii) general (policy-derived) measures. Changes in states and sub-states trigger the assignment of specific schedules. Further and different schedules can readily be implemented.

#### World initialization

Information about the world is georeferenced using geographical data retrieved from OpenStreetMap including building types and distances. Each building is considered a location as long as its floor area is large enough and it is not a non-residential building, e.g., a windmill. The building types are assigned according to labels in the input data. For example, buildings marked as ‘office’ or ‘industrial’ are assigned as workplaces while buildings marked as ‘public’ or ‘church’ are assigned as public places. All buildings that are not labeled specifically are assigned as residential buildings or homes. If the input data did not contain a location for a hospital or morgue, these locations are added artificially at the margin of the world, during the initialization process. The sets of building types and location assignments can be adapted to the requirements of a specific community. An initialized world can be stored and, thus, used for comparative simulation of different scenarios.

#### Initializing the agent population

Each residential building is home to one household that was randomly drawn from a distribution based on German census data. According to the respective household type and considering the reported demography, agents with adequate ages are randomly generated. Each agent is assigned a specific weekly schedule, that comprises times spent at home, work or school, public places etc., which are based on predefined flexible schedules for different age groups and types of individuals. These differ from individual to individual and for different days of the week. During the initialization, the status of a predefined number of agents, chosen from a minimal number of households, is set to infected (I). All other agents are defined as susceptible (S).

This spatio-temporal network, defined by periodically recurring movement patterns, constitutes the environment in which agents interact and, as an inevitable consequence, spread the infection. Agents operate in defined sub-networks, specified by regularly visited locations.

#### Spatio-temporal simulation and modeling the spreading of SARS-Cov-2 infection

During each time-step of the simulation (one hour), the agents visit the locations within the georeferenced network specified by their respective schedules. Each agent is considered to encounter interactions with other agents at these locations. The interaction partner is randomly picked among all other agents present at the same location at the same time. Given a susceptible agent (S) which has been assigned to an infected agent (I) or *vice versa* as interaction partner, the infection transmission is a situation-dependent property occurring with a specific probability, *P_infection_*, upon which the uninfected agent’s state transits from susceptible to infected (I). This infection probability reads

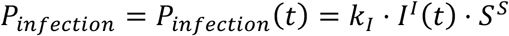

where *I^I^*(*t*) is the infectivity of an infected agent (I) depending on the duration of its infection and *S^s^*is the age-dependent susceptibility of a susceptible agent (S). *k_I_* is the infection rate, which represents individual protective measures of both parties (e.g. wearing face-masks) or location-specific factors such as the more strict hygiene regime in medical facilities.

#### State transitions of infected agents

Infected agents have a probability to change their status to either (R) or (D) or change their sub-status to (I^d^), (I^d^_H_) or (I^d^_ICU_). These probabilities are derived from published rates for those processes. The transition probabilities used to simulate the model are calculated for all transitions as probability per hour, where their dependence on age and on the duration of ongoing (sub-)states is taken into account.

#### Tracing of agent data from simulation

All locations and (sub-)states of individual agents are recorded and their time-course over the entire simulation is stored to be used as input for subsequent simulations and for analyses and plotting. Furthermore, information on the infection network (when, where and by whom every modelled agent was infected) can be extracted from a completed simulation. To test the effect of different interventions, parameter values, or the application and relaxation of social distancing, repeated simulations (with the different measures imposed) can be performed with identical initializations or starting from a selected time point of previous simulations to compare alternative trajectories of disease-spread.

### Model Analysis

The (instantaneous) effective reproduction number R_eff_ is defined as the average number of secondary infections caused by a single infected individual at a given time. The GERDA model calculates R_eff_ at time t as follows: for time t a backward-looking sliding time window is applied (window width used was 4 days) and all individuals that were infectious at some point within this window are identified. The ultimate number of secondary infections caused by these individuals was then averaged (including zeros for individuals not causing any secondary infections), yielding R_eff_(t).

### Data Sources

#### Johns Hopkins Coronavirus Resource Center

https://coronavirus.jhu.edu/map.html

#### Our World in Data

https://ourworldindata.org/coronavirus#all-charts-preview

#### Robert Koch Institute [RKI]

https://www.rki.de

1. Current Stage/Situation report of the Robert Koch Institute (RKI) about COVID-19:

1.1. COVID-19 fatalities reported to RKI sorted according to age and gender
1.2. Reported COVID-19 cases per 100.000 inhabitants in Germany sorted according to age groups and gender
1.3. Case numbers
1.4. Intensive care numbers / see Divi Intensive Register
2. SARS-CoV-2 Characteristics for Coronavirus-Disease-2019 (COVID-19) (in German) https://www.rki.de/DE/Content/InfAZ/N/Neuartiges_Coronavirus/Steckbrief.html Data from the characteristics refer to different studies
3. Modelling of exemplary scenarios of the SARS-CoV-2-epidemics 2020 in Germany https://edoc.rki.de/handle/176904/6547.2

#### Divi-Intensive register [DIVI]

https://www.divi.de

1. Daily reports of the DIVI Intensive register
2. Case numbers of reported intensive care patients (to estimate transition probabilities)

#### Federal Office of Statistics

Census, Distribution of household types, size and corresponding age distributions

1. https://ergebnisse.zensus2011.de/
2. https://service.destatis.de/

#### OpenStreetMap

Geospatial data for initialization of locations openstreetmap https://www.openstreetmap.de/

### Code Availability

The code will be available shortly at https://tbp-klipp.science/GERDA/code/

## Data Availability

The code will be available shortly at https://tbp-klipp.science/GERDA/code/

## Acknowledgements

This work was supported by the Deutsche Forschungsgemeinschaft (DFG: Cluster of Excellence MATH+, TRR 175) and by the German Ministry of Education and Research (BMBF, Liver Systems Medicine (LiSyM) network grant) and by the People Programme (Marie Skłodowska-Curie Actions) of the European Union’s Horizon 2020 Programme under REA grant agreement no. 813979 (‘Secreters’). Escalera-Fanjul X is supported with a postdoctoral grant from CONACYT (CVU 420248). We wish the populations of Gangelt, Heinsberg, Epping, and Vaxholm all the best and would like to point out that the GERDA model remains an abstraction and it is neither a repetition of history nor can it be traced back to real individuals, thus safe-guarding the privacy of the public.

## Author contributions

Conceived the project: RL, EK

Conceptual work on the model: BG, SOA, OB, JAHW, RL, EK

Programming: BG, SOA, OB, JAHW, AK, LB, JELH, MK^a^, RUTM, MS

Analysis and computational experiments: BG, SOA, OB, JAHW, AK, MK^a^, RUTM, MS, EK

Data collection: XEF, JELH, MK^b^,LM, HP, PSS

Wrote the paper: BG, OB, JAHW, MK^b^, MS, RL, EK

All authors agreed on the final version of the manuscript.

## Supplementary Information

The supplementary materials include:

**Supplementary Table 1:** Quantitative information used for state changes.

**Supplementary Fig. 1:** Age-dependent trajectories for S, I, R, and D.

**Supplementary Fig. 2:** Results for Heinsberg.

**Supplementary Fig. 3:** Variation of infectivity *k_I_*.

**Supplementary Fig. 4:** Timing of lockdown.

**Supplementary Fig. 5:** Variation of reopen times of schools after closure of all locations at 200h.

**Supplementary Fig. 6:** Variation of reopen times of all locations after closure of all locations at 200h.

**Supplementary Fig. 7:** Effect of non-compliance.

**Supplementary Fig. 8:** Re-infections or import of new infections after the infection wave.

**Supplementary Fig. 9:** Alternative scenario for comparison of tracing strategies.

**Supplementary Fig. 10:** Comparison of model-predictions with reported cases for the municipality of Gangelt.

**Supplementary Fig. 11:** Comparison of age group specific contact and infection frequencies for the municipality of Gangelt in baseline and lockdown scenarios.

**Supplementary Fig. 12:** Comparison of bimodal behavior for four different geo-locations with different population sizes.

**Supplementary Fig. 13:** Stringency of non-pharmaceutical interventions.

**Supplementary Fig. 14:** Age distribution.

**Supplementary Fig. 15:** Effect of lockdown time on number of deceased people.

**Supplementary Table 1:**
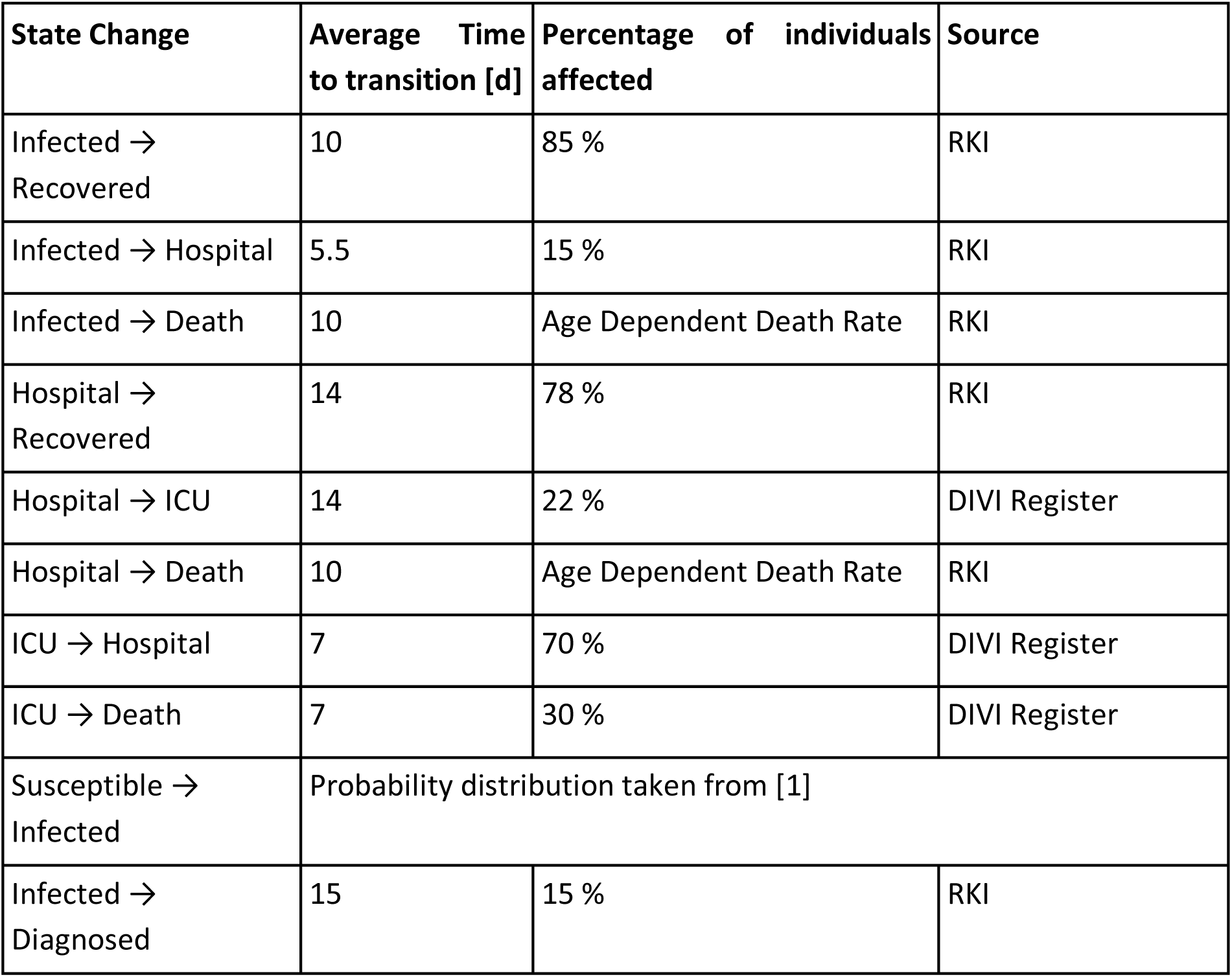
Quantitative information used for state changes. Our model utilizes transition probabilities for the propagation of SARS-CoV-2/COVID-19 [4]. For state changes in the left column we derived age-dependent probabilities per hour for the transition to occur from the following data: average time in a state until transition (given in days) and percentage of affected people undergoing transition. This information is intrinsically incomplete and ever evolving. For example, diagnostic capacity in Germany is currently increasing, hence, the respective parameters should be calibrated during the pandemic or for new geolocations. The infection probability by itself is hard to determine as statistics for SARS-CoV-2 infection events are incomplete. Thus, these values are educated guesses challenged with sensitivity analysis (**Supplementary Fig. 3-7**).

| State Change | Average Time to transition [d] | Percentage of individuals affected | Source |
| --- | --- | --- | --- |
| Infected → Recovered | 10 | 85 % | RKI |
| Infected → Hospital | 5.5 | 15 % | RKI |
| Infected → Death | 10 | Age Dependent Death Rate | RKI |
| Hospital → Recovered | 14 | 78 % | RKI |
| Hospital → ICU | 14 | 22 % | DIVI Register |
| Hospital → Death | 10 | Age Dependent Death Rate | RKI |
| ICU → Hospital | 7 | 70 % | DIVI Register |
| ICU → Death | 7 | 30 % | DIVI Register |
| Susceptible → Infected | Probability distribution taken from [1] |  |  |
| Infected → Diagnosed | 15 | 15 % | RKI |

**Supplementary Fig. 1:**
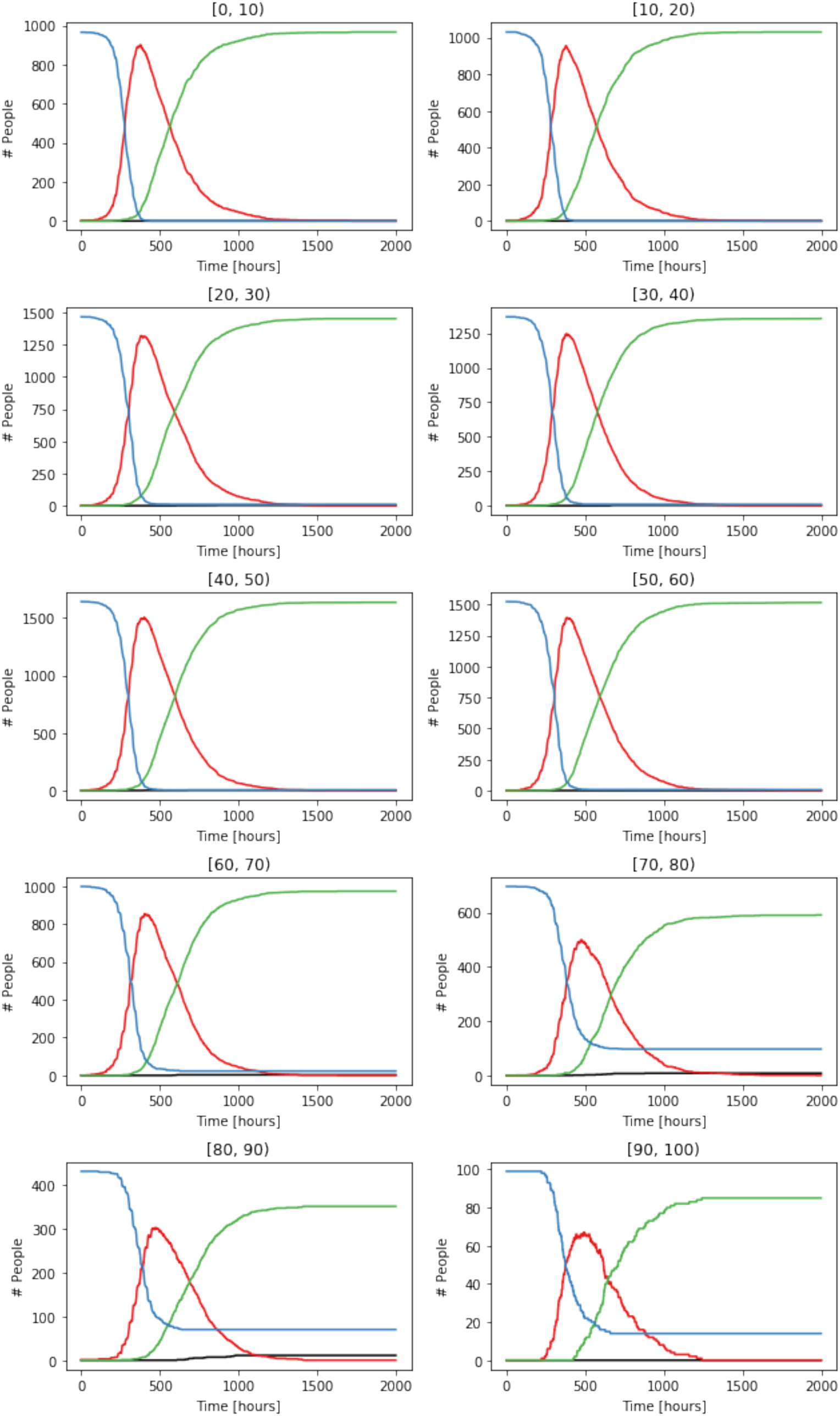
Age-dependent trajectories for S, I, R, and D. The individual curves show results for a single simulation for infection rate 0.3. Age ranges are given on top of each panel.

**Supplementary Fig. 2:**
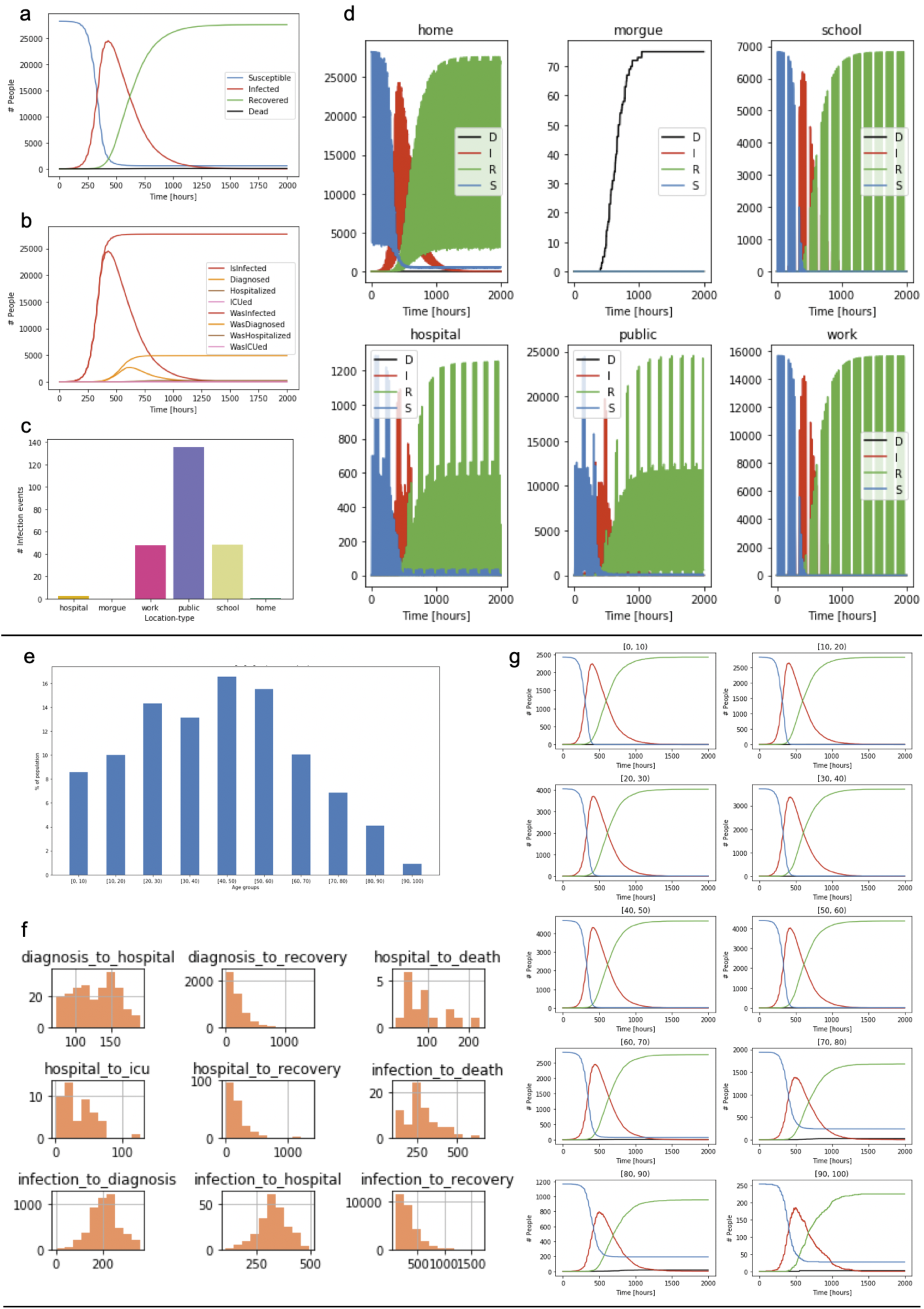
Results for Heinsberg. We have also initialized the model with geographical demographic data for Heinsberg, Gangelt’s larger neighbor town with about 28.300 inhabitants. **a**, Time courses of agents’ state (S), (I), (R), and (D). **b**, Time courses of sub-states of infection. **c**, Dynamics at different location types. **e**, Age distribution. **f**, Distribution of status dwelling times before a specific status transition. **g** Status trajectories as in a for age groups.

**Supplementary Fig. 3:**
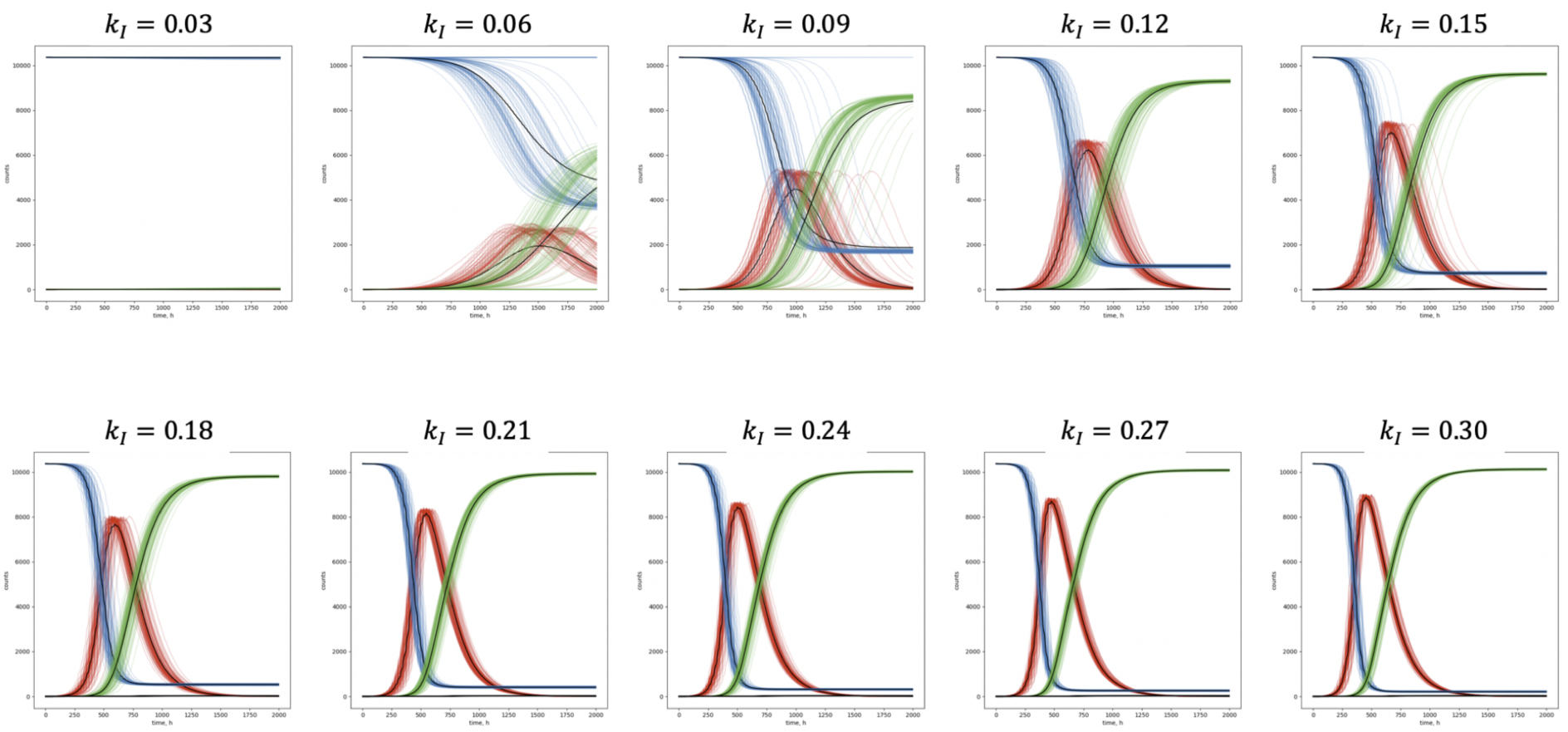
Variation of infectivity. ***k_I_***. The simulation is performed for different values of infectivity and no interventions imposed.

**Supplementary Fig. 4:**
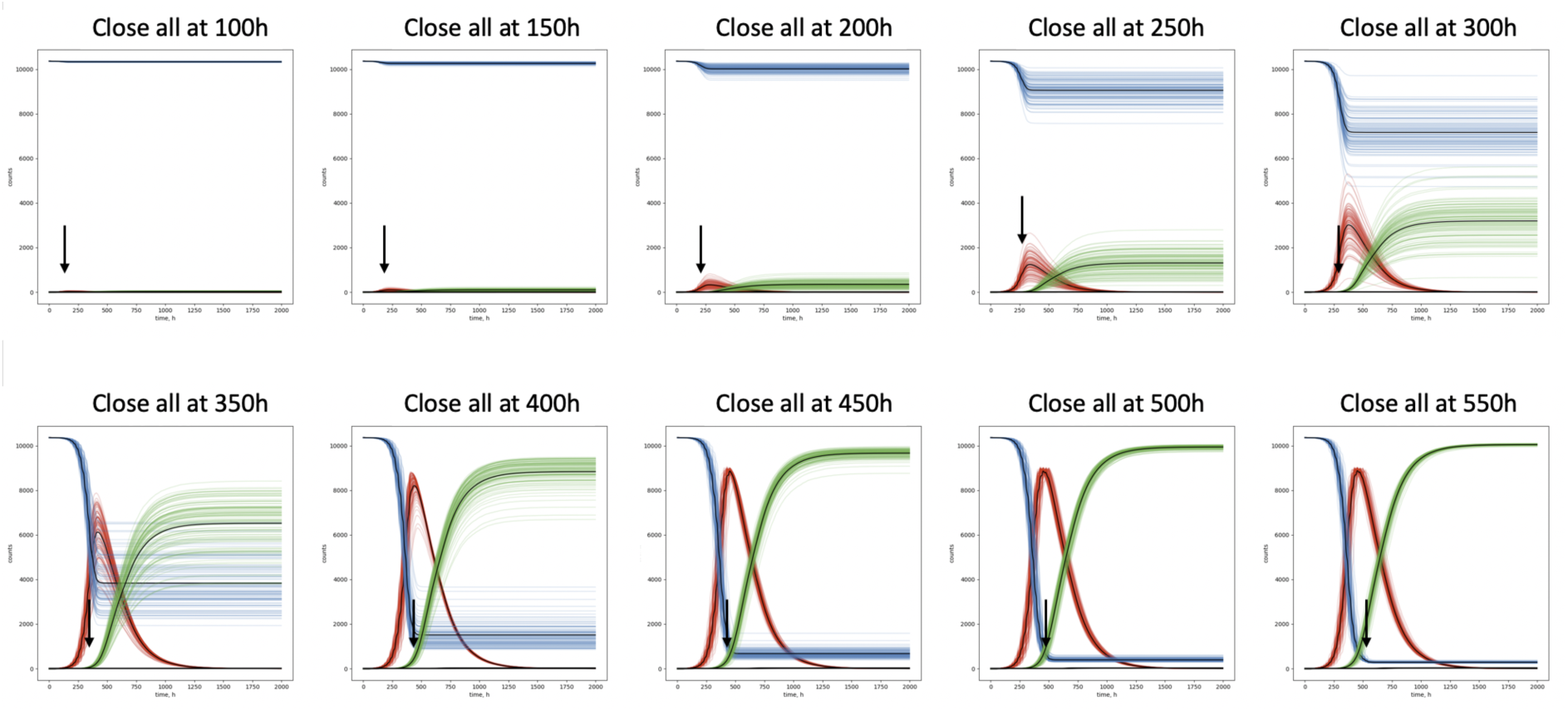
Timing of lockdown. The effect of different starting times to realize the general measure of lockdown, i.e. closure of all facilities.

**Supplementary Fig. 5:**
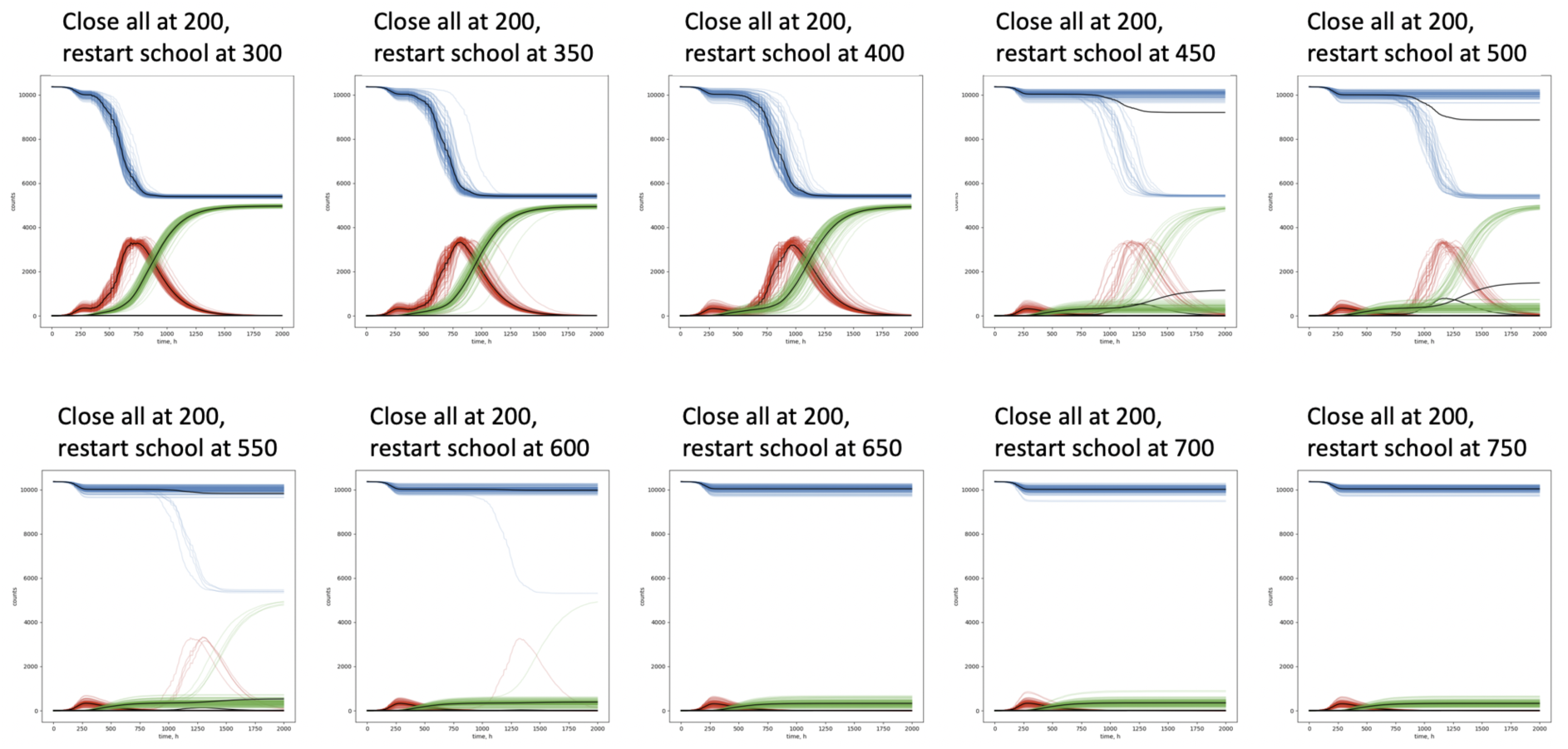
Variation of reopen times of schools after closure of all locations at 200h. Parameter value *k_inf_ = 0.3*.

**Supplementary Fig. 6:**
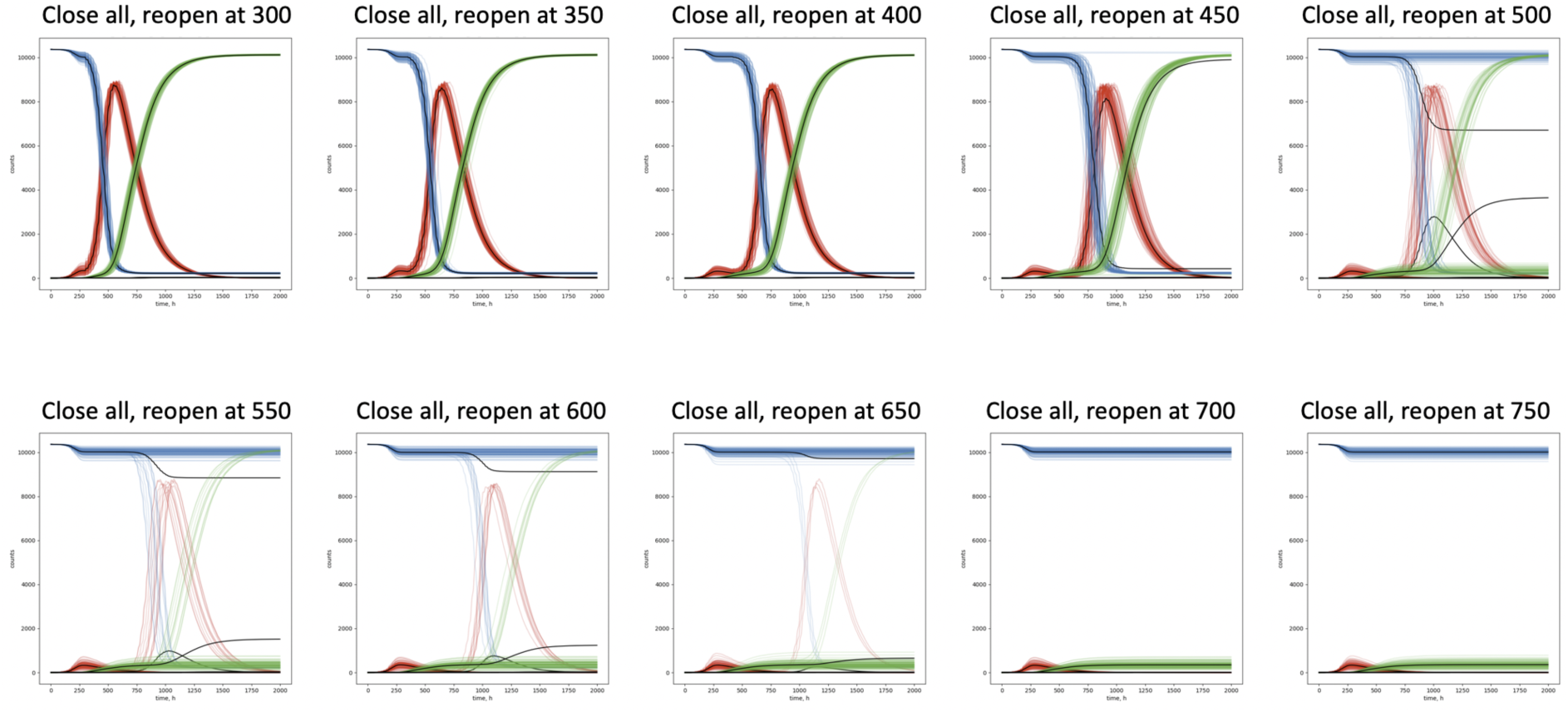
Variation of reopen times of all locations after closure of all locations at 200h. Parameter value *k_inf_ = 0.3*.

**Supplementary Fig. 7:**
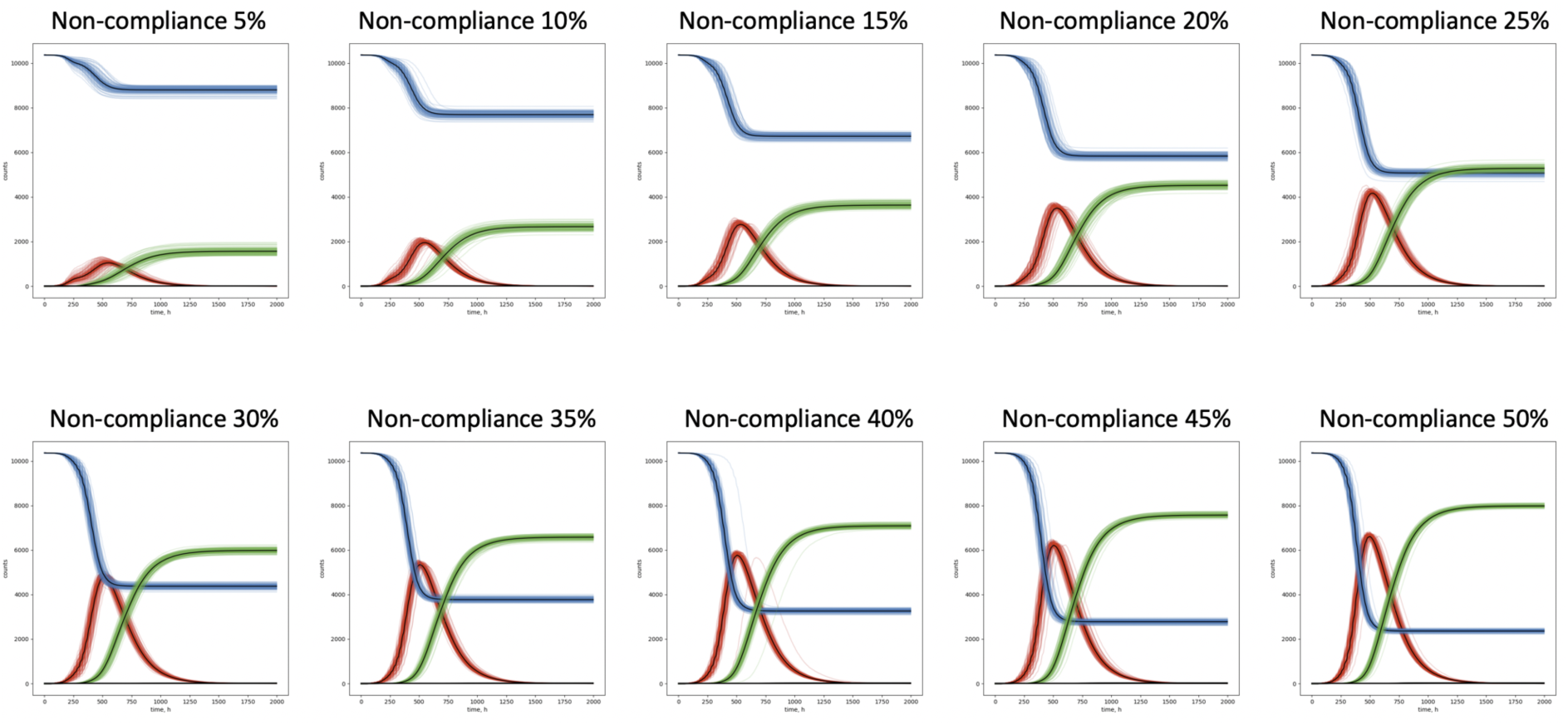
Effect of non-compliance. This scenario presents a full lockdown (closure of schools, work and public) at 200h. The percentage given in each panel indicates the percentage of individuals who are not following, either due to lack of willingness or since they have duties such as system-relevant work.

**Supplementary Fig. 8:**
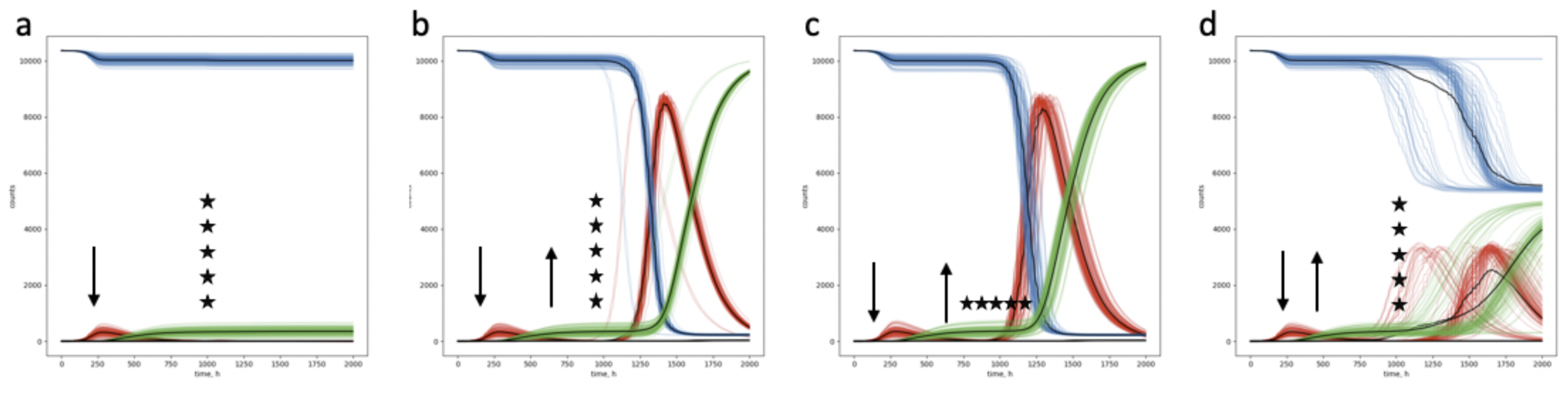
Re-infections or import of new infections after the infection wave. All scenarios: close all at 200h, **a**, 5 re-infections at 1000h. **b**, Reopen all at 750h, 5 re-infections at 1000h, **c**, Reopen all at 750h, re-infections at 800h, 900h, 1000h, 1100h, 1200h, 1300h (1 each time). **d**, Reopen school at 750h, 5 re-infections at 1000h. All panels: arrow down close all, arrow up reopening, stars indicate the entry of new infectees from outside.

**Supplementary Fig. 9:**
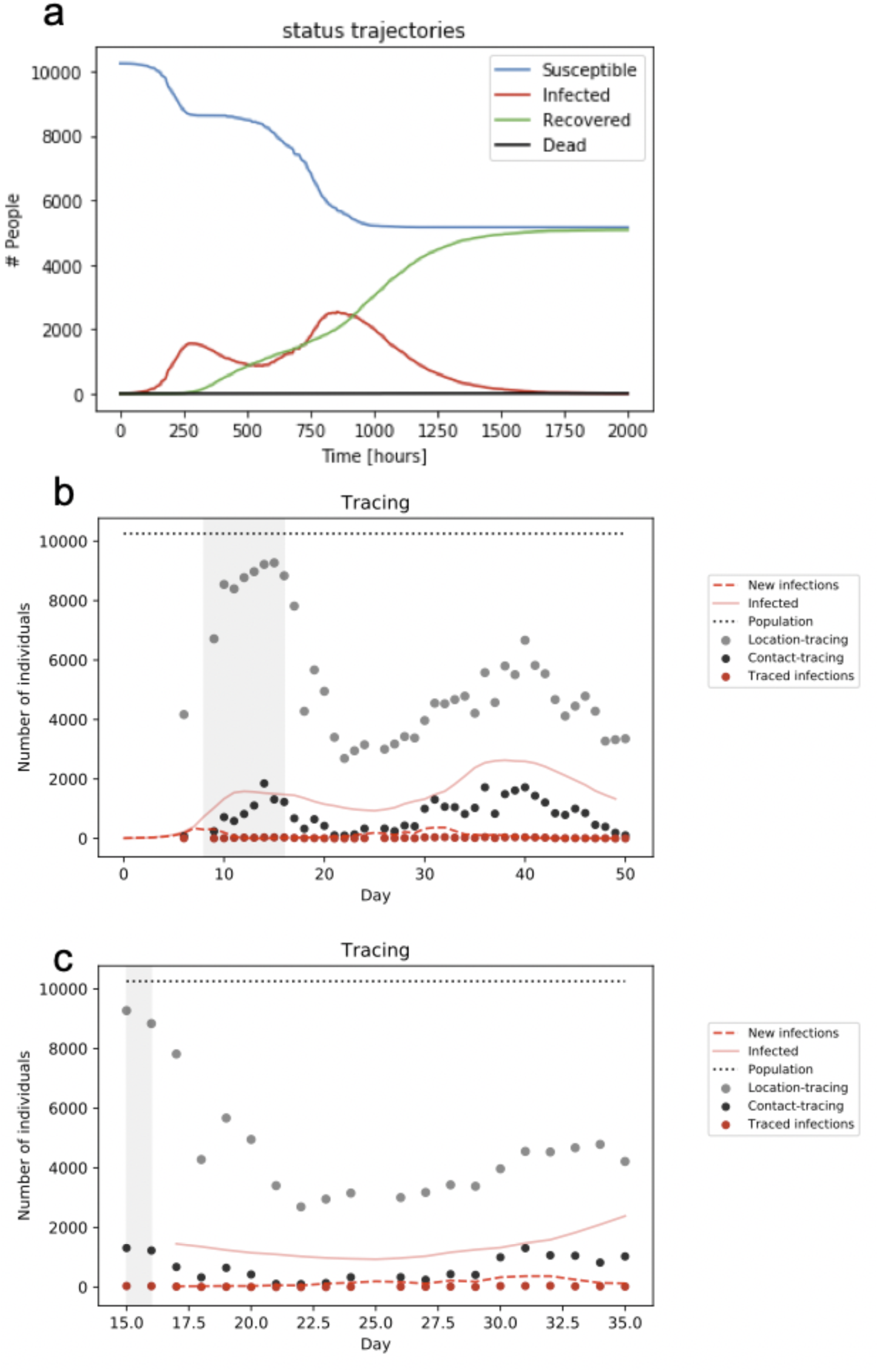
Alternative scenario for comparison of tracing strategies. a. Trajectories for a lockdown at 200h and full reopening at 400h. **b**, Time courses of infections that occurred and that could be location or contact traced. Shaded grey area indicates the period of the lockdown. It is apparent that during the lockdown, newly diagnosed cases still result in potentially infectious contacts to be traced. **c**, As in **b**, magnifying the relevant time window following the 20 days after the exit from lockdown.

**Supplementary Fig. 10:**
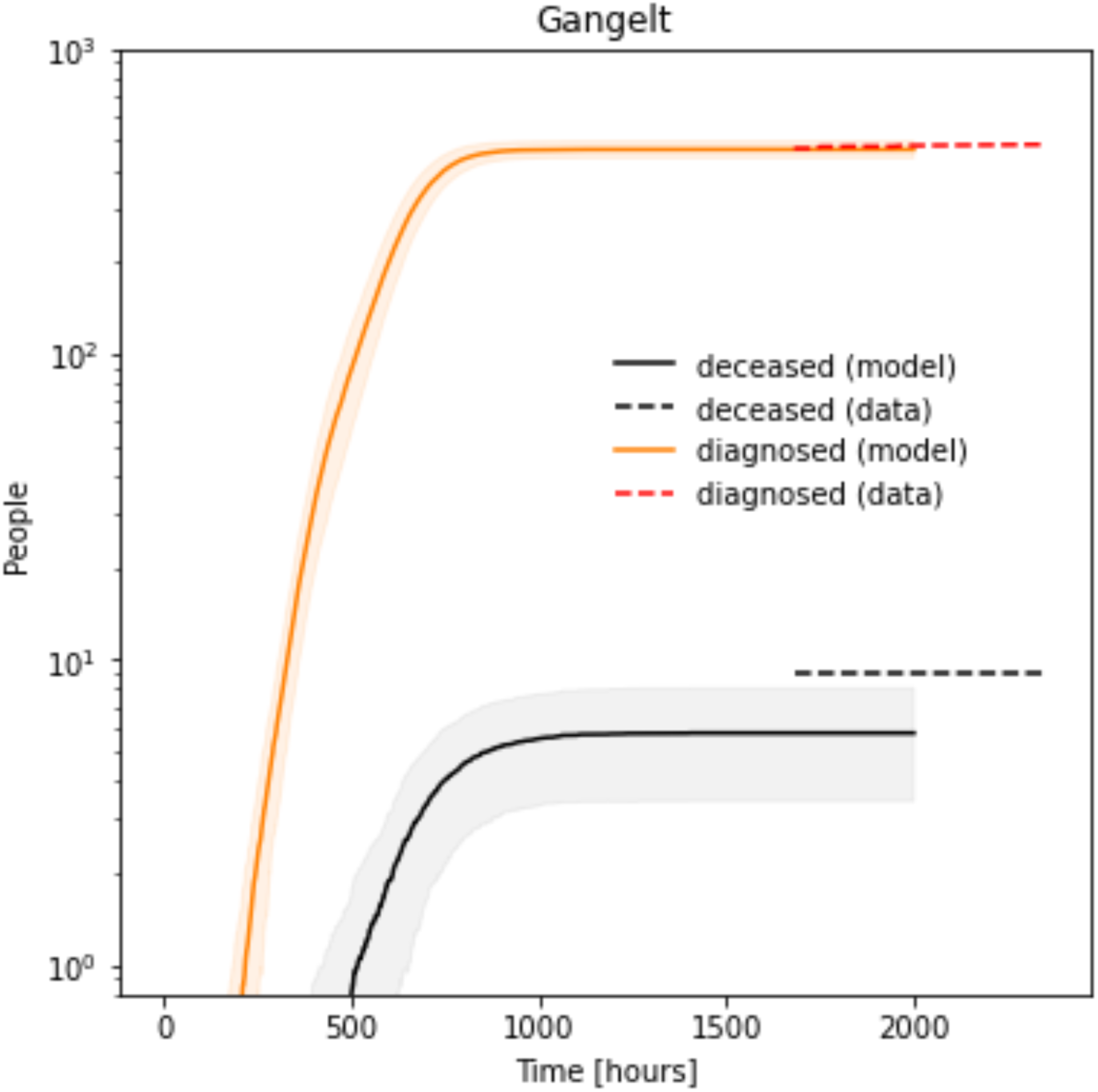
Comparison of model-predictions with reported cases for the municipality of Gangelt. The number of diagnosed cases for the municipality of Gangelt (orange), predicted by the model (mean over 100 replicate simulations with standard deviation indicated by shaded area) over the period from 15. February 2020 (carnival event *‘Kappensitzung’*to the 8th May 2020. The dashed line indicated the reported (diagnosed) infections between the 24th of April and the 22nd of May 2020. The number of deaths (black), predicted by the model in the same period with the reported number of deceased individuals, indicated by the dashed line.

**Supplementary Fig. 11:**
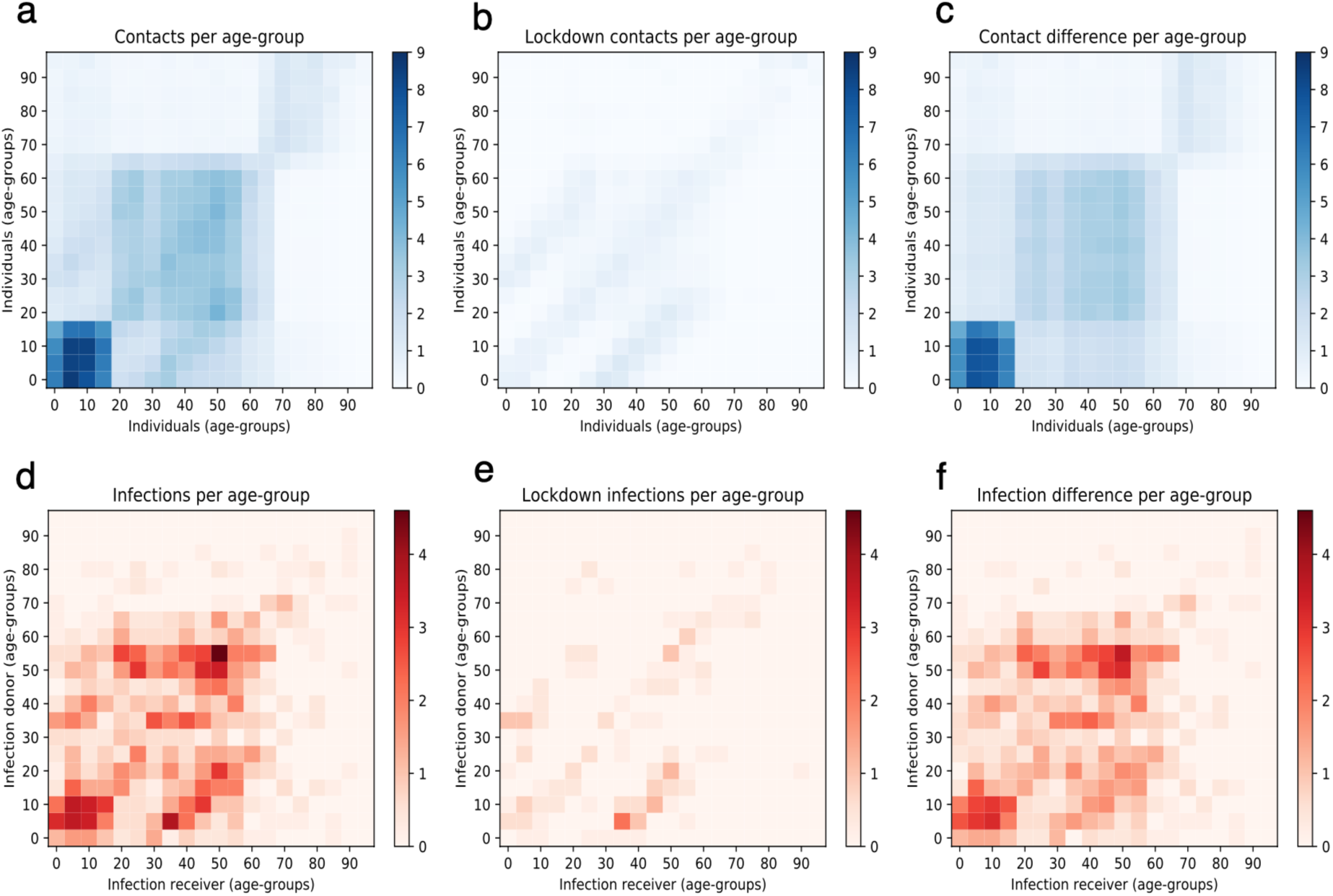
Comparison of age group specific contact and infection frequencies for the municipality of Gangelt in baseline and lockdown scenarios. a. Average number of daily unique contacts between members of different age groups in the baseline scenario over a period of 5 successive working days. **b**, Average number of daily unique contacts between members of different age groups during lockdown. **c**, Difference in contacts between baseline and lockdown scenarios, i.e. contacts prevented by the lockdown. **d**, Average number of infections per day, between members of different age groups in the baseline scenario over a period of 5 working days. **e**, Average number of infections per day between members of different age-groups during lockdown. **f**, Difference in infections between baseline and lockdown scenarios, i.e. infections prevented by the lockdown.

**Supplementary Fig. 12:**
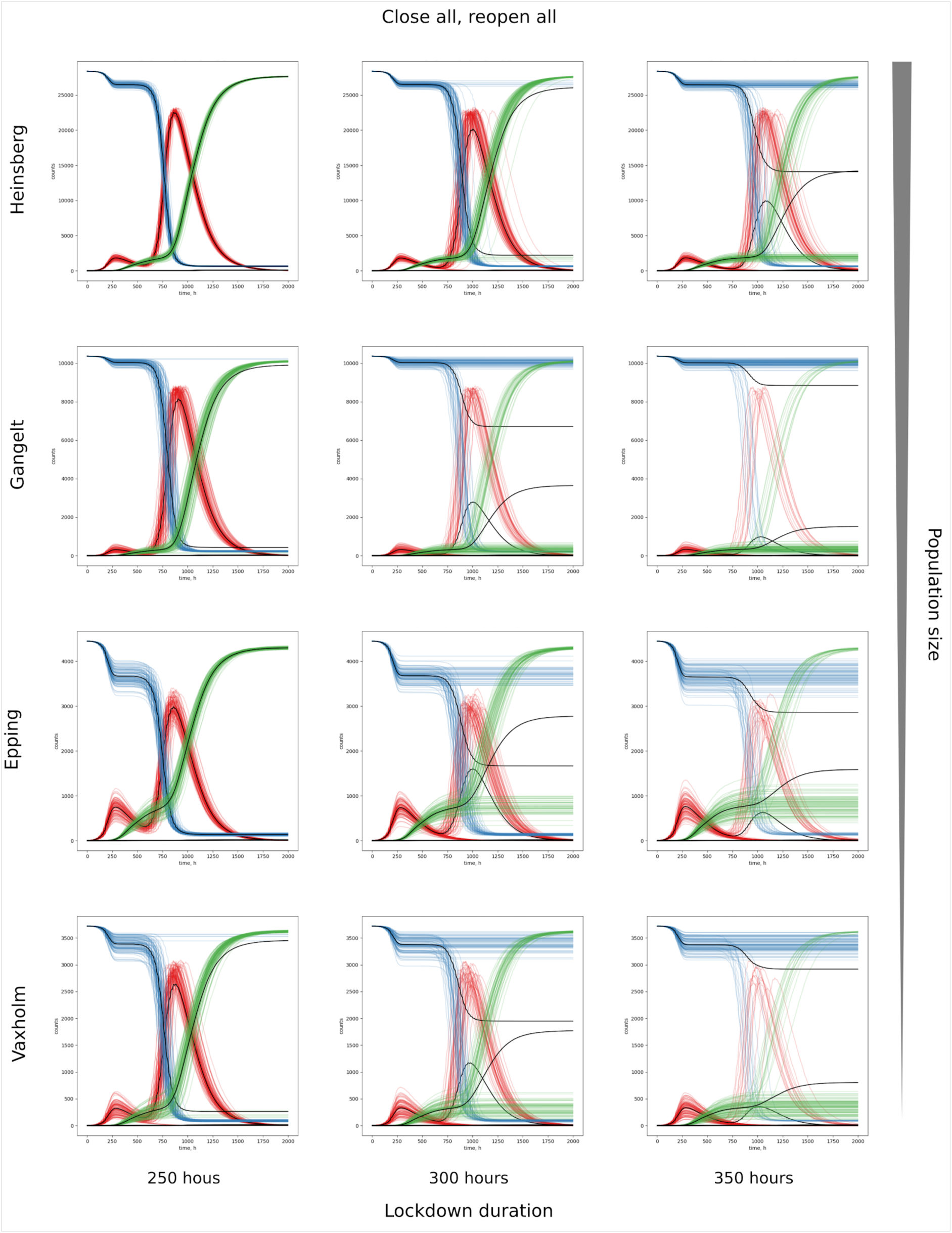
Comparison of bimodal behavior for four different geo-locations with different population sizes. We simulated lockdown and reopen scenarios for the four different communities: Heinsberg (Germany), Gangelt (Germany), Epping (UK) and Vaxholm (Sweden) starting with 5 infected individuals. Here we find bimodal behavior for all four geo-locations depending on the lockdown duration.

**Supplementary Fig. 13:**
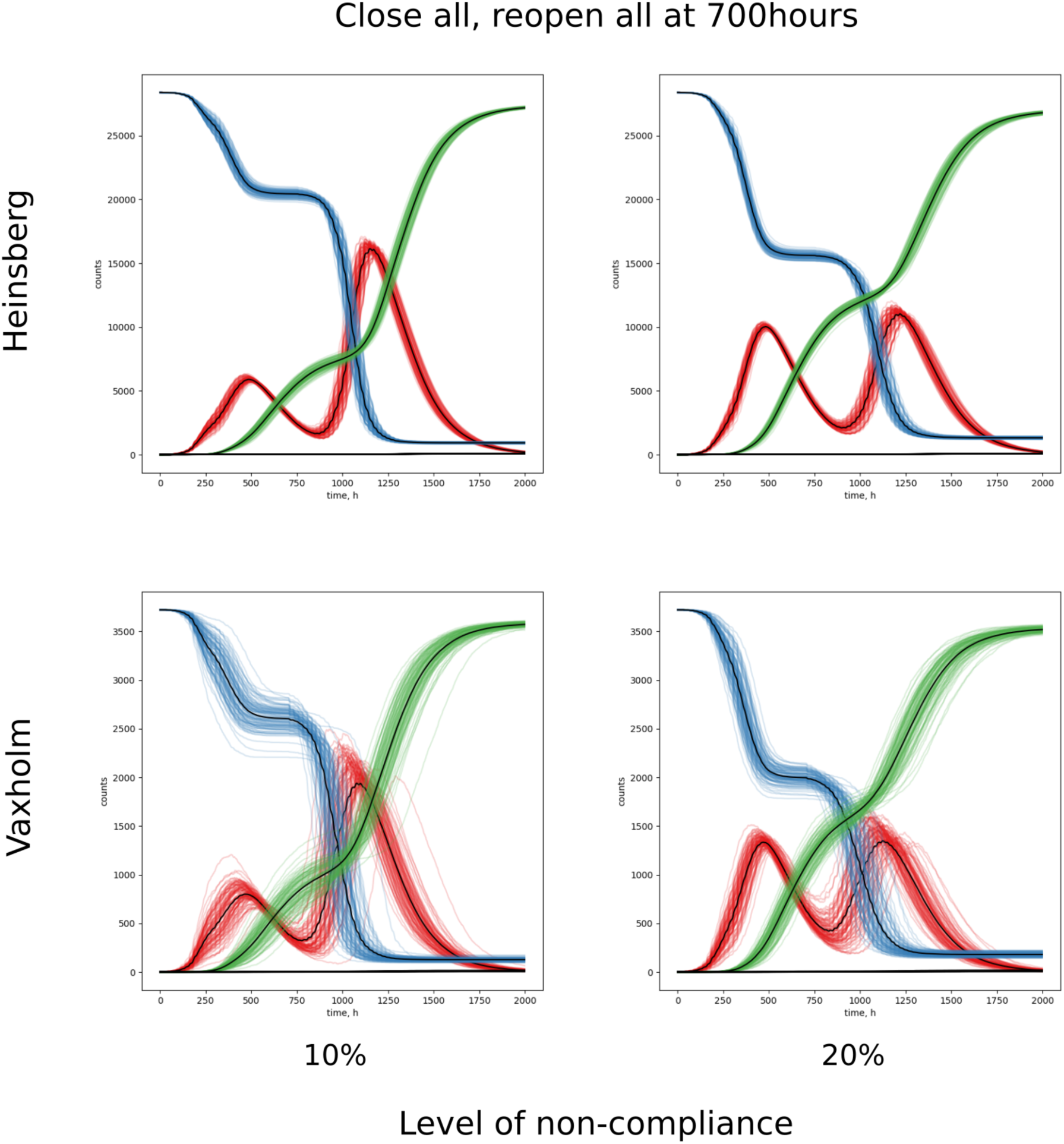
Stringency of non-pharmaceutical interventions. Since complete lockdown is practically hard to achieve in real-world societies, we simulated various levels of non-compliance (**Supplementary Fig. 7** Here, we compare the effect of the level of non-compliance to lockdown (at 200h) on the size of a second peak after reopening (at 700h). We see for these examples that non-compliance of 10% leads to a smaller first peak with more remaining susceptibles and a higher second peak than non-compliance of 20% where both peaks in infected people are of roughly the same height. Depending on the capacity of health care systems, the height of the second peak can be critical for the ability to cope with the threat or breakdown.

**Supplementary Fig. 14:**
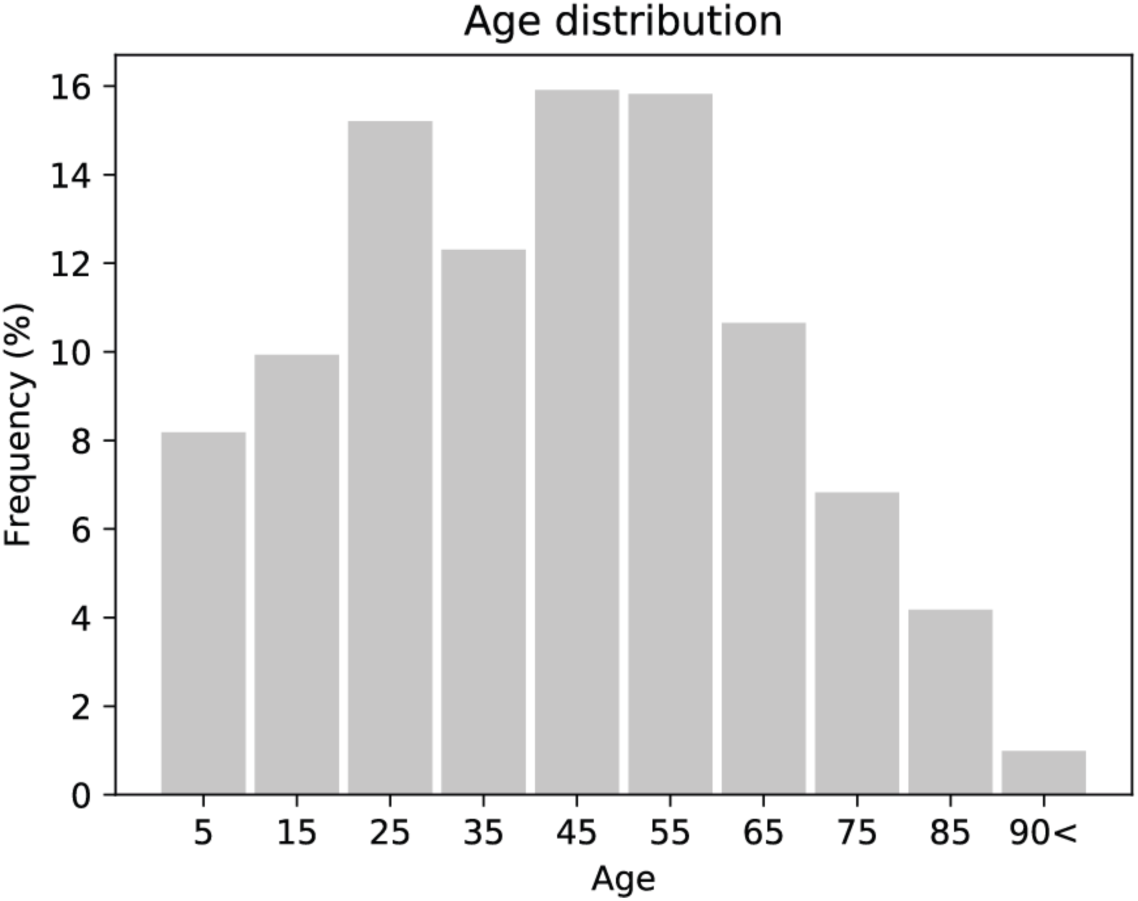
Age distribution. Resulting age distribution for Gangelt.

**Supplementary Fig. 15:**
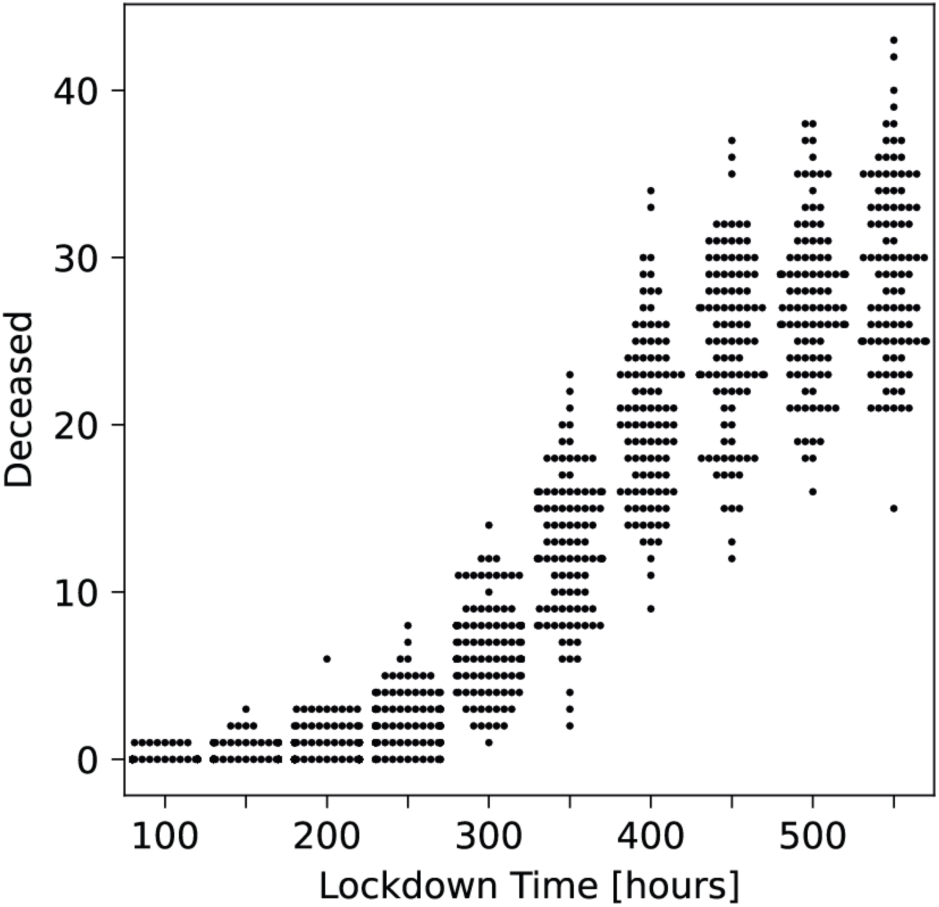
Effect of lockdown time on number of deceased people. Distribution of the total number of deceased in 100 simulations for different lockdown starting times.

